# Network medicine-based epistasis detection in complex diseases: ready for quantum computing

**DOI:** 10.1101/2023.11.07.23298205

**Authors:** Markus Hoffmann, Julian M. Poschenrieder, Massimiliano Incudini, Sylvie Baier, Amelie Fitz, Andreas Maier, Michael Hartung, Christian Hoffmann, Nico Trummer, Klaudia Adamowicz, Mario Picciani, Evelyn Scheibling, Maximilian V. Harl, Ingmar Lesch, Hunor Frey, Simon Kayser, Paul Wissenberg, Leon Schwartz, Leon Hafner, Aakriti Acharya, Lena Hackl, Gordon Grabert, Sung-Gwon Lee, Gyuhyeok Cho, Matthew Cloward, Jakub Jankowski, Hye Kyung Lee, Olga Tsoy, Nina Wenke, Anders Gorm Pedersen, Klaus Bønnelykke, Antonio Mandarino, Federico Melograna, Laura Schulz, Héctor Climente-Gonzalez, Mathias Wilhelm, Luigi Iapichino, Lars Wienbrandt, David Ellinghaus, Kristel Van Steen, Michele Grossi, Priscilla A. Furth, Lothar Hennighausen, Alessandra Di Pierro, Jan Baumbach, Tim Kacprowski, Markus List, David B. Blumenthal

**Author notes:** The authors wish to be known that, in their opinion, the first eight authors should be considered as shared first authors (Markus Hoffmann, Julian M. Poschenrieder, Massimiliano Incudini, Sylvie Baier, Amelie Fitz, Andreas Maier, Michael Hartung, Christian Hoffmann). The authors wish to be known that, in their opinion, the last seven authors should be considered as shared last authors (Priscilla A. Furth, Lothar Hennighausen, Alessandra Di Pierro, Jan Baumbach, Markus List, David B. Blumenthal).

## Abstract

Most heritable diseases are polygenic. To comprehend the underlying genetic architecture, it is crucial to discover the clinically relevant epistatic interactions (EIs) between genomic single nucleotide polymorphisms (SNPs)^1–3^. Existing statistical computational methods for EI detection are mostly limited to pairs of SNPs due to the combinatorial explosion of higher-order EIs. With NeEDL (**ne**twork-based **e**pistasis **d**etection via **l**ocal search), we leverage network medicine to inform the selection of EIs that are an order of magnitude more statistically significant compared to existing tools and consist, on average, of five SNPs. We further show that this computationally demanding task can be substantially accelerated once quantum computing hardware becomes available. We apply NeEDL to eight different diseases and discover genes (affected by EIs of SNPs) that are partly known to affect the disease, additionally, these results are reproducible across independent cohorts. EIs for these eight diseases can be interactively explored in the Epistasis Disease Atlas (https://epistasis-disease-atlas.com). In summary, NeEDL is the first application that demonstrates the potential of seamlessly integrated quantum computing techniques to accelerate biomedical research. Our network medicine approach detects higher-order EIs with unprecedented statistical and biological evidence, yielding unique insights into polygenic diseases and providing a basis for the development of improved risk scores and combination therapies.

## Introduction

Genome-wide association studies (GWAS) aim to identify genetic single nucleotide polymorphisms (SNPs) that are individually associated with a phenotype such as a disease^1–3^. Thousands of individual SNPs have been associated with diseases since the early 1990s, yet they account only for a fraction of the investigated traits’ heritability^4, 5^. The hypothesis is that a significant proportion of the missing heritability can be explained by epistatic SNP interactions that are jointly but not individually associated with the phenotype^6^. However, no undisputed cases of epistasis in humans are known. Therefore, scalable detection tools for epistatic interactions that yield interpretable, high-quality results are needed to advance the understanding of possible genetic causes of diseases.

Detecting biologically plausible epistatic candidatė SNP sets is difficult: Firstly, there is no consensus on the choice of the formal model in order to make this problem algorithmically accessible. Secondly, it is often unclear if predicted cases of epistasis are statistical artifacts since they are hardly interpretable. Thirdly, comprehensively testing higher-order interactions is computationally intractable due to the combinatorial explosion of the search space. Existing epistasis detection tools, including Potpourri^7^, LinDen^8^, PoCos^9^, MACOED^10^, and BiologicalEpistasis^11^ hence do not scale to large datasets and are mostly restricted to pairwise interactions (Suppl. Table 10). Finally, easily accessible resources to browse and explore pre-computed epistasis candidates interactively are lacking. To overcome these challenges, we present NeEDL (**ne**twork-based **e**pistasis **d**etection via **l**ocal search), an epistasis detection tool leveraging quantum computing and network medicine to identify biologically interpretable candidate sets of epistatic interaction between SNPs (see Figure 1-Figure 2). NeEDL unifies various GWAS input formats and initially filters datasets for relevant SNPs (Suppl. Fig. 1a). NeEDL then offers different statistical epistasis models^12^ for associating higher-order SNP sets with phenotypes, prioritizing SNP sets using a biologically-informed SNP-SNP interaction (SSI) network. Our hypothesis is that SNPs affecting the same or functionally related proteins are more likely to be involved in relevant epistatic interaction and less likely to be statistical artifacts. Thus, SNPs are mapped to proteins using dbSNP^13^ and connected if they affect the same protein or a functionally associated protein (e.g., in a protein-protein interaction (PPI) network, Figure 2a). Within the SSI network, NeEDL uses local search with multi-start and simulated annealing to find connected subgraphs of a user-specified size that are locally optimal w. r. t. the selected statistical epistasis model (Figure 2b). Focusing on SNP sets inducing connected subgraphs in the SSI network not only increases the likelihood of uncovering biologically meaningful cases of epistasis but also dramatically reduces the size of the search space. Finally, NeEDL fully integrates quantum computing (QC) algorithms to yield high-quality initial solutions for the local search to reduce NeEDL’s runtime further once QC hardware becomes more readily available. To the best of our knowledge, NeEDL is the first application to seamlessly integrate QC for solving a real-world life sciences problem.

**Figure 1.**
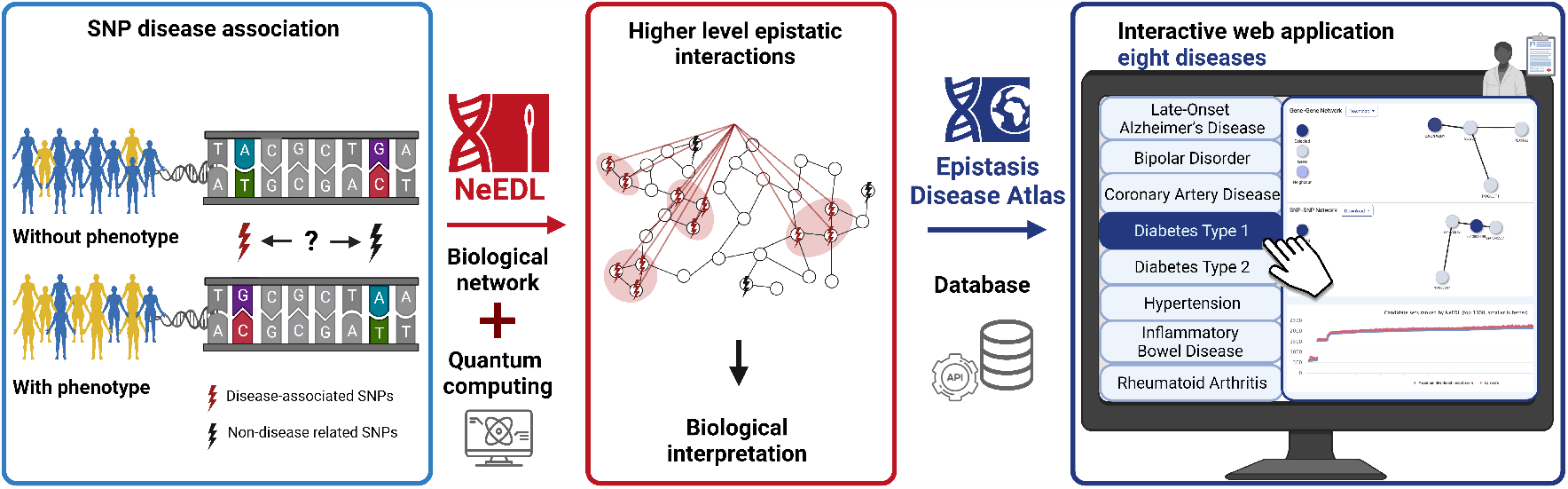
Overview of the study. Genome-wide association studies (GWAS) have been used to identify single nucleotide polymorphisms (SNPs), which are specific positions in the genome where a single nucleotide substitution occurs and are prevalent in a significant portion of the population. These SNPs serve as markers to locate and identify regions of the genome that may be responsible for common complex diseases. In the presented cohorts, individuals displaying the disease phenotype that is associated with SNPs are highlighted in yellow. The prediction of disease phenotypes often involves the combined influence of multiple SNPs, as the individual effects of a single SNP may not be sufficient to manifest the disease phenotype. NeEDL (**ne**twork-based **e**pistasis **d**etection via **l**ocal search) is a novel epistasis detection tool that harnesses the strengths of biological networks and quantum computing to tackle the combinatorial explosion challenge, making the study of higher level epistatic interactions more feasible. By employing a network medicine approach, NeEDL narrows the search space to expedite the analysis of higher-order genetic interactions in complex traits. As a final step, the most promising SNP candidates for eight diseases are made available in a database for interactive exploration via the web-based Epistasis Disease Atlas

**Figure 2.**
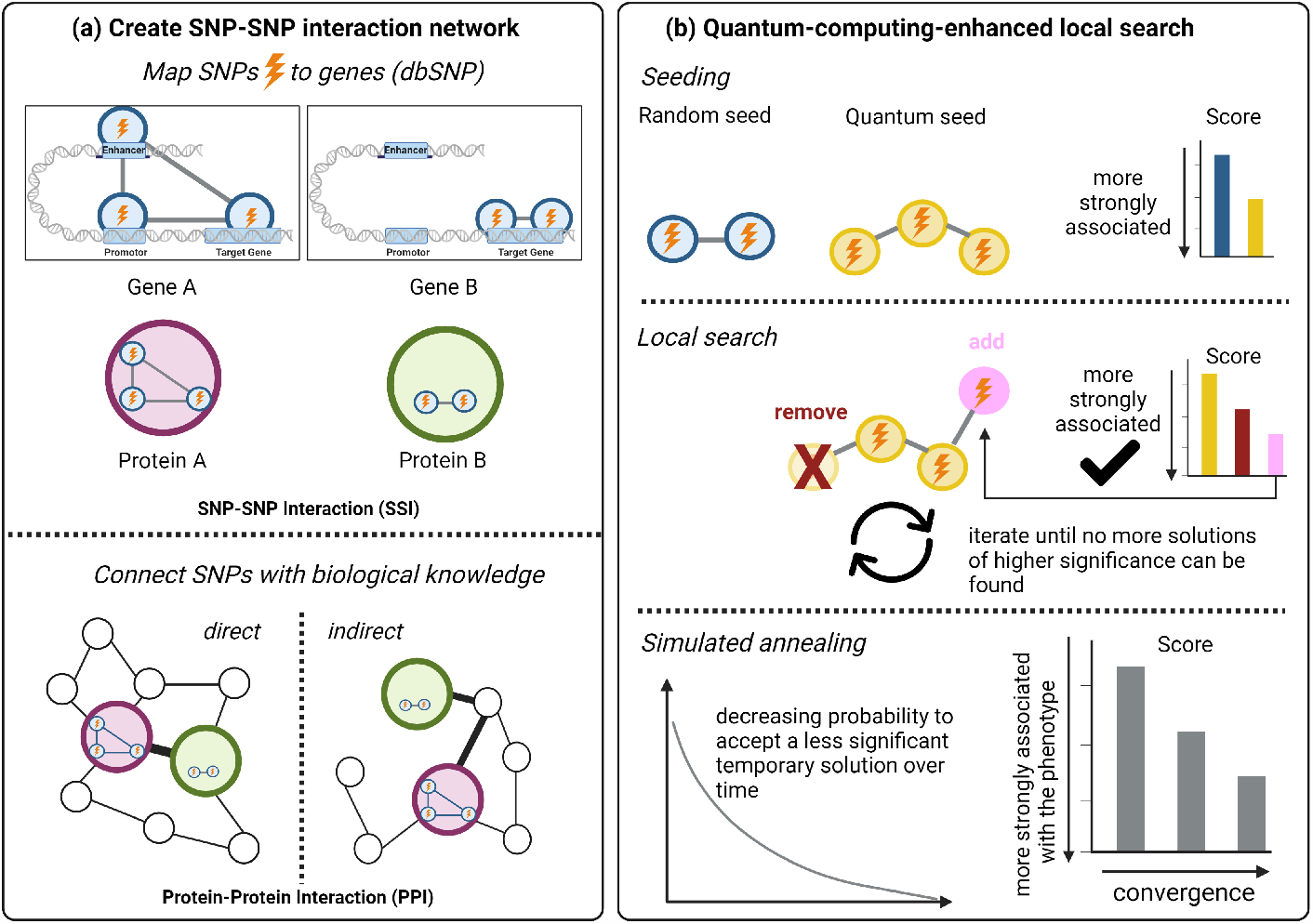
Overview of the methodology. (a) SNPs are mapped to proteins via SNP-gene links obtained from dbSNP^13^ and are then connected in an SNP-SNP interaction (SSI) network if they are mapped to the same protein or to adjacent proteins in a protein-protein interaction (PPI) network (i. e., directly when the proteins interact with each other or indirectly connected when the SNPs are linked through other proteins). (b) Next, NeEDL picks either random seeds or seeds selected by a quantum-computing optimization algorithm. It then uses local search (i.e., either adding new SNPs from the direct neighborhood in the SSI network or removing SNPs that worsens the score) to find SNP sets of a user-specified maximum size that induce connected subgraphs in the SSI network and are locally optimal w. r. t. statistical association of the higher-order genotype with the investigated phenotypic trait. To quantify association strength, NeEDL implements various statistical epistasis models suggested in the literature^12^. Since local search can get stuck due to the requirement of constant improvement in each step, we use simulated annealing, which allows to accept a less significant intermediate solution with decreasing probability over time.

We benchmark NeEDL against frequently used epistasis detection tools on GWAS data for eight human diseases — Late-Onset Alzheimer’s Disease (LOAD), Bipolar Disorder (BD), Coronary Artery Disease (CAD), Diabetes Type 1 (T1D), Diabetes Type 2 (T2D), Hypertension (HT), Inflammatory Bowel Disease (IBD), and Rheumatoid Arthritis (RA) (see Suppl. Table 1-9 for an overview over sample numbers and SNP numbers and Datasets for information on datasets). Our validation shows that NeEDL markedly outperforms the currently used epistasis detection tools in terms of the discovered candidate SNP sets’ associations with the diseases (i.e., the statistical score). The top-scoring candidate SNPs sets are further supported by (i) significant biological associations with the respective diseases, and (ii) in the case of LOAD, by replication in independent cohorts (i.e., discovery with NeEDL in the WTCCC cohort and replication in the UK Biobank cohort). We make the results of NeEDL for the eight diseases, which we obtained in over one million CPU hours, widely accessible through the Epistasis Disease Atlas (https://epistasis-disease-atlas.com, Suppl. Fig. 1b) via an application programming interface (API), an R and Python package, an FTP server, and an interactive, feature-rich web application.

## Results

### NeEDL outperforms existing epistasis detection tools

We compared NeEDL to the existing epistasis detection tools LinDen^8^ and MACOED^10^ on the same input data as NeEDL. All tools were run with default hyper-parameters (see Methods). PoCos^9^, Potpourri^7^, and BiologicalEpistasis^11^ were excluded due to unresolvable implementation problems and/or excessive runtime or memory requirements (Suppl. Table 11). To ensure fair competition, we compared the SNP sets using four widely used scores in this research area^12^, including the *P* -value of the *χ*^2^-test (optimized for by LinDen), the Bayesian network score K2 (optimized for by MACOED), the negative log-likelihood score of the maximum likelihood model (MLM) (optimized for by NeEDL per default) and the differences between the negative log-likelihoods of fitted linear and quadratic regression models (NLL gain)^12^. For the *P* -value of the *χ*^2^-test, the Bayesian network score K2, and the MLM score, low scores indicate promising SNP sets. Conversely, the higher the NLL gain, the more promising the scored SNP set (see Methods). To show the robustness of the results and to control if higher-order candidate SNP sets achieve more significant associations just because of the number of SNPs, we generated baselines by randomly sampling 1,000 size-matched SNP sets (size 2 for LinDen and MACOED, higher-order for NeEDL) to estimate the false positive rate (see Methods). All *P* -values (both for the background distributions and for the results) were adjusted using the Benjamini-Hochberg procedure^14^.

Since the local search algorithm returns a result for each start seed, we obtain thousands of results. To find a suitable cut-off, we show the MLM and K2 scores for the 100 top-ranked SNP sets computed by NeEDL (Figure 3a, Suppl. Fig. 2b, Suppl. Fig. 3). Interestingly, we observe phase transitions (i.e., an abrupt change to a lower significance of the score over linear increasing ranks) for both scores, which typically occur after 50 SNP sets (except for the LOAD and IBD datasets, with transitions between the ranks 20 and 30, and the RA dataset, with transitions at 60 SNP sets). We hypothesize that this score gap represents the barrier between promising and not promising epistasis candidate sets and thus focused on the top 50 candidates (i. e., where most transitions occur) selected based on the respective metric each tool is optimizing for (NeEDL: MLM score, LinDen: *P* -value of *χ*^2^-test, MACOED: K2 score). MACOED was executed 100 times to account for randomized subroutines, leading to the union of the top 50 candidate SNP sets for each run for MACOED. The data depicted in Figure 3b (Suppl. Fig. 2b, Suppl. File 1), demonstrates that NeEDL outperforms LinDen, MACOED, and the baselines across all metrics and datasets. Even in instances where it performs comparably on some metrics on certain datasets, NeEDL still surpasses the competitor on at least one other metric across all datasets in terms of statistical association. LinDen, as well as the second-order- and higher-order baseline, are outperformed on all metrics over all datasets. On all datasets, the *χ*^2^-test *P* -values obtained for the SNP sets computed by NeEDL approach the maximum number of decimal places available in standard computation. With the more complex MLM and K2 scores, differences become more visible, and we see that NeEDL markedly outperforms MACOED in all datasets (Figure 3b, Suppl. Fig. 2b). It is noteworthy that when the cut-off of the rank is set at 25 for LOAD, and IBD and 60 for RA, the advantage of NeEDL over MACOED becomes more distinct (Suppl. Fig. 4). NeEDL also yields significant lower K2 scores than MACOED on all datasets, even though NeEDL did not optimize for this score. Since NeEDL optimizes for the MLM score, we can see that NeEDL outperforms all competitors and baselines on this score. In terms of NLL gain, NeEDL again outperforms its competitors on all datasets except IBD, where the NLL gain is similar to MACOED. Also, for LOAD, very low values of the NLL gain in MACOED indicate that, unlike NeEDL, MACOED mainly finds SNP sets with strong additive main effects. In terms of required computational resources, NeEDL and LinDen (both executable on desktop PCs) are much more efficient than MACOED (which requires a super-computer with high RAM; Suppl. Table 12). Figure 3c and Suppl. Fig. 2c shows that in most datasets, the best scores are achieved with three to seven interacting SNPs. Since we ran NeEDL with a maximum SNP set size of 10, NeEDL likely does not exploit combinatorial statistical artifacts but prioritizes SNPs that may be involved in protein-protein interactions and thus be more likely to be biologically meaningful. We examined whether the use of the biological priors encoded in the SSI network indeed guides NeEDL toward more promising SNP sets. For this, we generated 100 randomized networks with preserved topology (by shuffling the node labels) and 100 randomized networks where the SNPs’ node degrees are preserved in expectation (Methods, Suppl. Fig. 5a). For this analysis, we randomly selected three of the eight datasets (BD, T2D, and RA) to avoid excessive runtime. MLM scores on the original SSI networks are, on average, better than those on the randomized networks (Figure 3d, Suppl. Fig. 2d).

**Figure 3.**
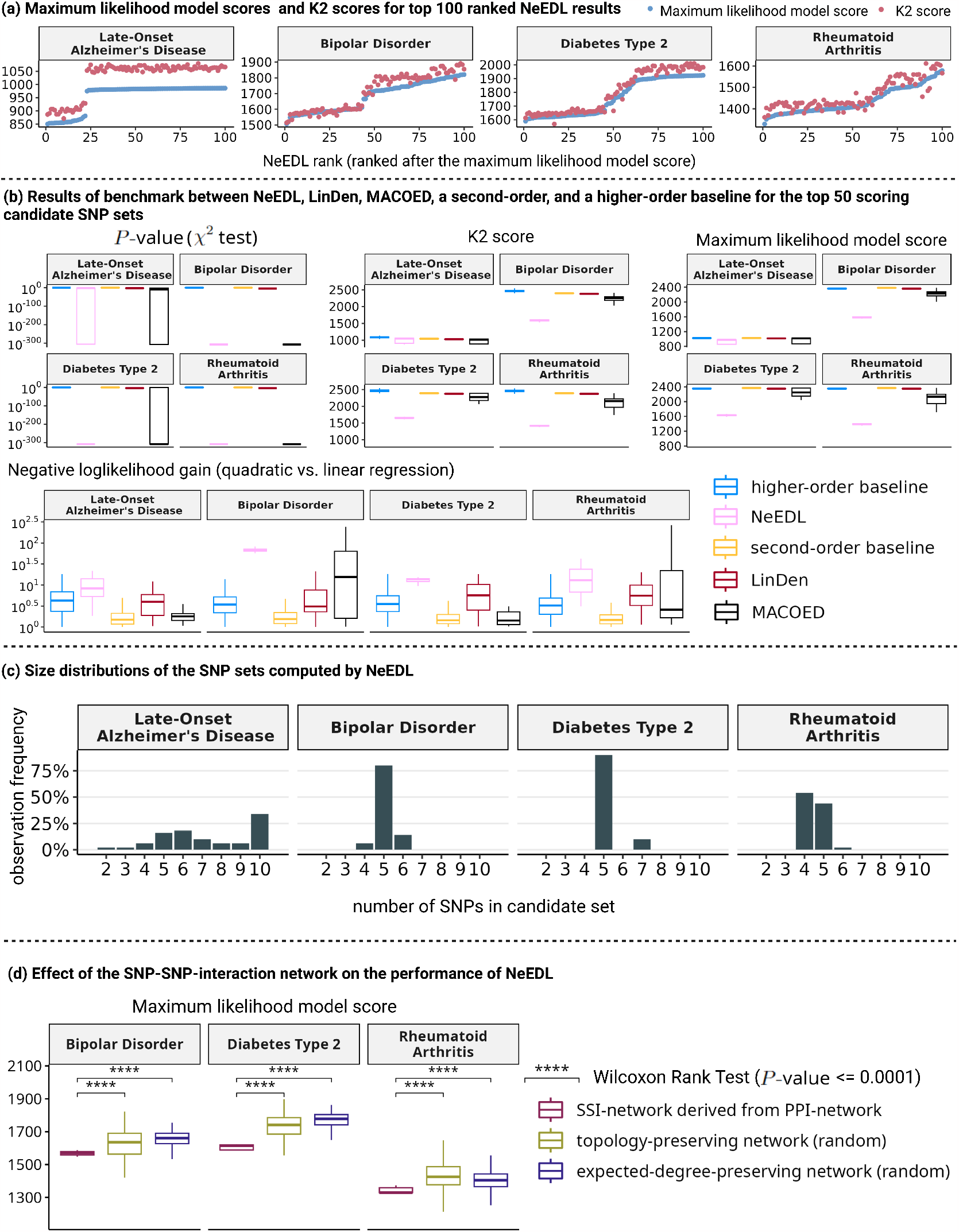
Quantitative evaluation of the SNP sets computed by NeEDL for Late-Onset Alzheimer’s Disease, Bipolar Disorder, Diabetes Type 2, and Rheumatoid Arthritis. Results for Coronary Artery Disease, Diabetes Type 1, Hypertension, and Inflammatory Bowel Disease can be found in Suppl. Fig. 2. (a) Visualization of the maximum likelihood model score and the K2 score of the top 100 candidate SNP sets ranked by NeEDL. (b) A benchmark study between NeEDL, LinDen, MACOED, a second-order baseline (i.e., 1000 random sampled pairs of SNPs), and a higher-order baseline (i.e., 1000 randomly sampled sets consisting of multiple SNPs) shows that NeEDL outperforms in statistical significance existing epistasis detection tools w. r. t. four different evaluation metrics. (c) Analyzing the number of SNPs included in NeEDL’s output, SNP sets reveal that the most promising SNP sets are typical of sizes between three to seven. (d) Comparing maximum likelihood model scores of NeEDL results against those obtained using randomized networks demonstrates that the use of the SSI network indeed leads to the discovery of more promising SNP sets.

### Consistent results across independent cohorts

We conducted a replication study to validate the findings of NeEDL and to demonstrate the consistency of the results across independent datasets for LOAD. For this, we retrieved two independent replication datasets (independent from the discovery dataset used in the benchmark) from the UK Biobank (see Methods), one with individuals of British ancestry (similar to the LOAD dataset) and one with individuals of mixed ancestry (i.e., 10% of the samples are of mixed ancestries like Latino, African, or Asian) to estimate population-specific effects. From the SNP sets computed by NeEDL on the LOAD dataset (i. e., the dataset used for discovery), we selected the 50 SNP sets with the most significant MLM scores (to be consistent with the benchmark) and re-computed their *χ*^2^-test *P* -values, MLM scores, K2 scores, and NLL gains.

Figure 4a shows that we could replicate the phase transition observed in the LOAD dataset used for the benchmark (Figure 3a): The top 22 SNP sets and the top 23 SNP sets of the replication data set of British ancestry and mixed ancestry, respectively, replicate with a significant *χ*^2^-test *P* -values below 0.05 (Suppl. Fig. 6). The MLM scores of replication correlate (Pearson correlation of 0.995) highly with the MLM scores of the benchmark study (Figure 4b). We can further observe a drop in the Pearson correlation to 0.993 in the mixed ancestry dataset, suggesting that the population-specific genetic architecture might play a role in epistasis detection. Similar to the LOAD dataset, a phase transition occurs in *P* -values and MLM scores after the top 22 SNP sets (Suppl. Fig. 6). We also re-computed *χ*^2^-test *P* -values, MLM scores, and K2 scores for the 50 best SNP sets computed by MACOED and LinDen on the LOAD benchmark dataset. Consistent with the discovery study, NeEDL with its top 50 candidate sets outperforms LinDen in both replication datasets (Figure 4c). MACOED is outperformed in the *P* -values of the *χ*^2^-test, the K2 score, and the MLM score. However, MACOED yielded similar results or slightly less signficiant scores with the NLL-gain. Overall, the replication results are consistent with the results of the benchmark in the discovery dataset.

**Figure 4.**
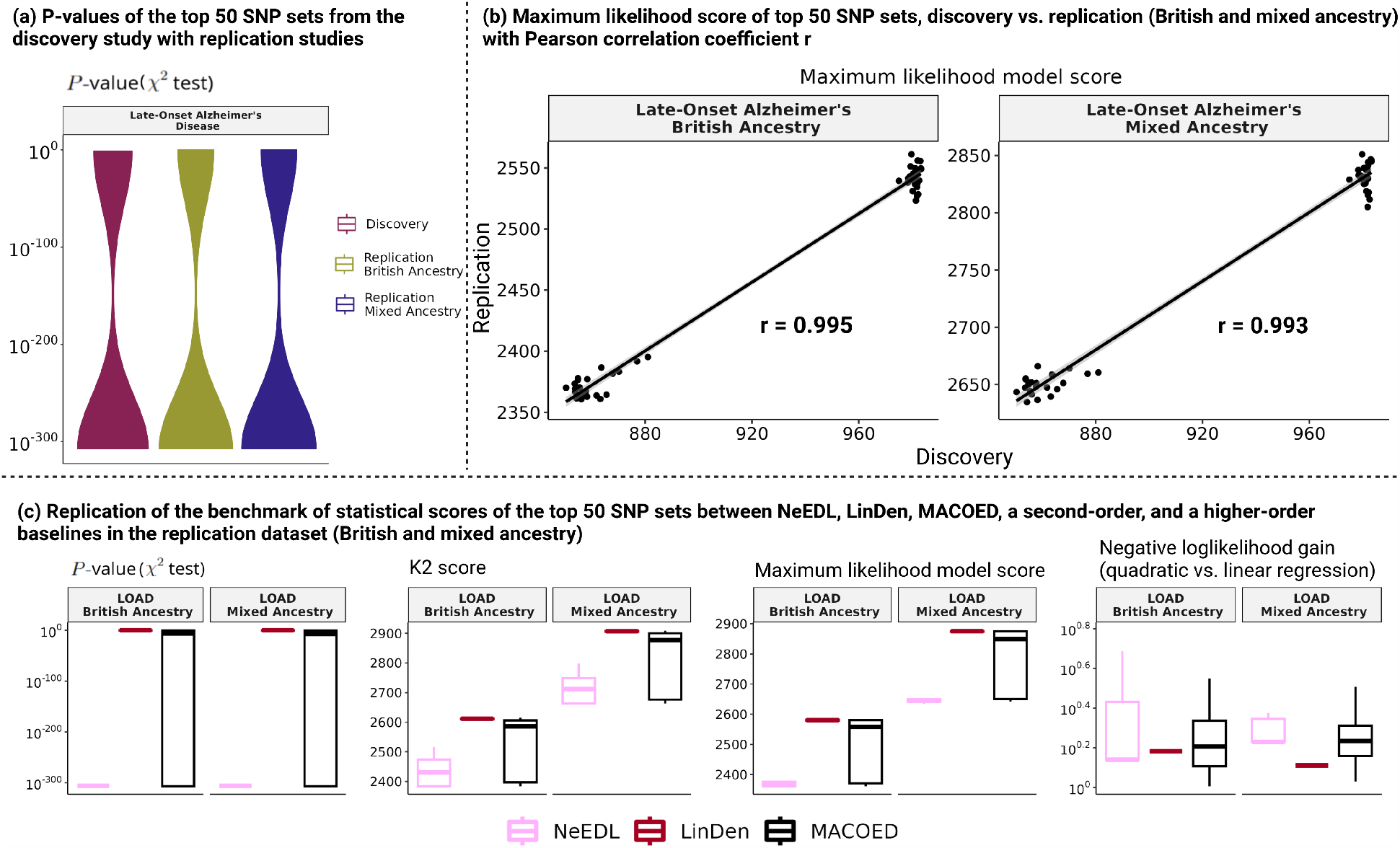
Replication studies in two independent data sets from UK Biobank with British and mixed ancestry. (a) *P* -values across discovery and the two replication studies. (b) Correlation of the MLM score between the discovery study and the two replication studies. (c) Replication of the benchmarking between NeEDL, LinDen, MACOED, a second-order baseline, and a higher-order baseline in two replication data sets.

#### Epistasis candidates uncovered by NeEDL are biologically plausible

By utilizing NeEDL, we identified potential epistatic candidate SNP sets that have been selected based on their maximum likelihood model scores. We selected LOAD, T1D, and IBD (the latter two in Suppl. Materials 1) as they are well-represented in the literature. In each of these diseases, various SNPS of a single gene in different combinations with SNPs of several other genes is present in the majority of the statistically most significant candidate SNP sets (Figure 4b, Suppl. Fig. 6). We evaluated the top genes before the gap that were affected by the predicted SNP interactions by performing gene set enrichment analysis (GSEA)^15, 16^ to identify statistically significant enrichment in Gene Ontology^15^ and KEGG^17^ (see Methods for details).

APOE was present in the 25 highest penetrance SNP combinations for the disease LOAD. APOE and the lipoprotein metabolism pathway have been previously recognized as the key gene for Alzheimer’s disease risk^18, 19^. Since APOE mutation status alone is not a reliable predictor of Alzheimer’s disease, a better understanding of epistatic interactions may be instrumental to develop better diagnostics. NeEDL identified 47 additional genes exhibiting potential epistatic SNP combinations with APOE (Suppl. File 2). GSEA determined that 15 genes (APOE, SORL1, SMAD3, LRP2, YBX3, TP63, PRKCB, DAB1, LRP1, IFI16, UBQLN1, A2M, DYRK1A, HMGXB4, and TRAF3IP1), including APOE, were enriched in a gene set that includes the process to reduce the response to the stimulus (GO:0048585; k/K=0.0088; *P* -value=6.25*e*^*−*11^; FDR *q*-value=9.82*e*^*−*7^). Of note, one of the epistatic SNP combinations identified by NeEDL was rs429358 and rs7412 in the APOE gene, which has been previously identified as a potential epistatic SNP combination^20^.

In the Human Protein Atlas^21^, we can observe that the identified genes are significantly expressed in tissues that are reported to be affected by LOAD, T1D, or IBD (Suppl. Fig. 7)^22–34^.

### Quantum computing improves seeding of local search

The GWAS datasets employed in this study cover up to 140,000 SNPs due to necessary cleaning and filtering steps (Suppl. Table 1-10), a small fraction of the 84.7 million SNPs^35^ in the human genome. It is hence likely that, in the future, we will see a sharp increase in the number of covered and mappable SNPs. In the NeEDL workflow, this would result in an SSI network containing millions of SNPs. To explore such a huge network via randomly seeded local search with reasonable coverage, we would have to dramatically increase the number of initial solutions — certainly, beyond the capacities of classical high-performance computing clusters. We thus explored the use of quantum computing in NeEDL (see Methods for details) to find a promising set of initial solutions for the local search instead of starting with random seeds as in the classical implementation (Suppl. Fig. 5b). To be able to use a quantum computer, we modeled the problem of finding promising initial solutions as a variant of the max-clique problem. This problem can be transformed into a Quadratic Binary Unconstrained Optimization (QUBO) problem^36^, which is then solved using several quantum computing machines and simulators, including the quantum annealer D-Wave Advantage 6.1, the superconducting-based IBM Perth, and IBM Lagos devices.

To test this approach, we subsampled our BD, RA, and T2D datasets to 100, 500, and 1000 SNPs and ran NeEDL with random and quantum-computing-based seeding. Running quantum-computing-based seeding on the entire datasets is still infeasible due to limitations of current quantum computing hardware. Suppl. Fig. 8 shows that, using quantum computing, fewer but higher-quality candidate SNP sets are returned, and that the speed up these high-quality initial solutions induce in the downstream local search outweighs the additional runtime needed for the quantum-computing-based seeding. Since quantum computing hardware cannot yet process big data, we estimate the speed up on realistically sized GWAS data across different devices (Figure 5). Notably, any quantum device, whether simulated or executed on actual hardware, scales better than the baseline classical algorithm. We may hence anticipate quantum computation to surpass the classical baseline in the near future, showcasing a potential quantum advantage, even though currently only small subproblems can be addressed. To conduct these experiments, we used more than 4.1 million seconds of computing time on quantum resources.

**Figure 5.**
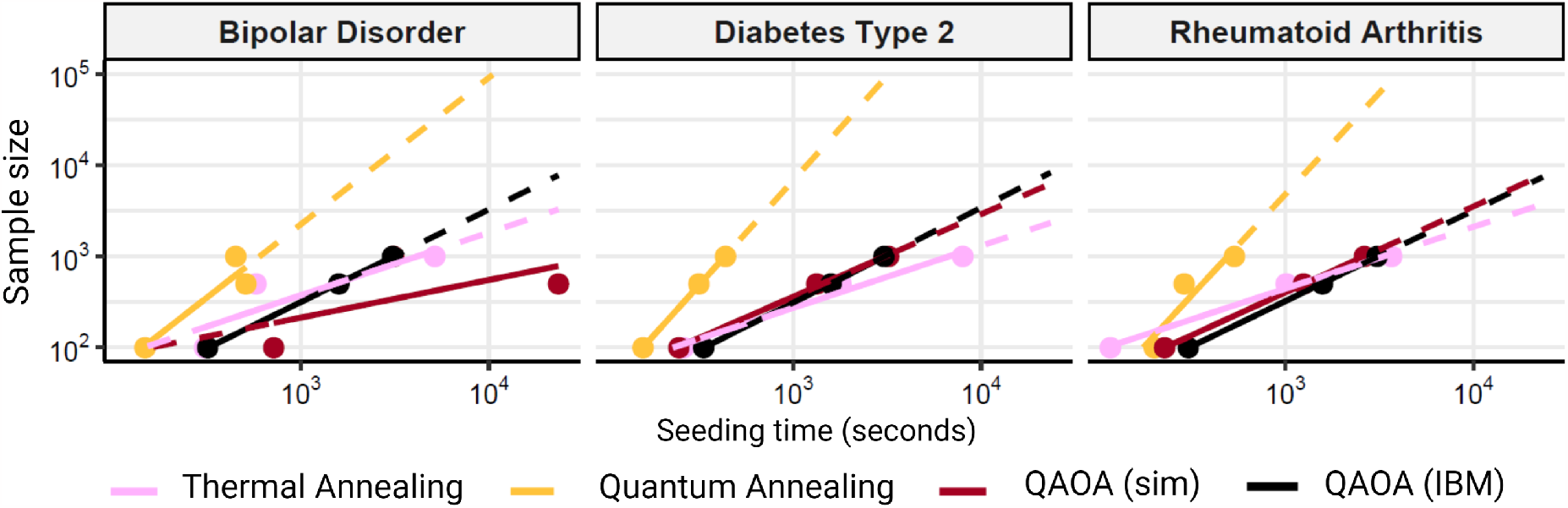
Experiments on the quantum computers: Expected scaling of the quantum hardware. The different quantum and classical devices used to perform the optimization process show different scaling, with the quantum devices (Quantum annealing, QAOA) having a higher slope than classical devices (Thermal annealing), suggesting a possible scaling advantage.

### Exploring epistatic interactions in the Epistasis Disease Atlas

Since more than one million CPU hours were necessary to calculate the results reported here, we make them widely and easily accessible through the web-based Epistasis Disease Atlas (https://epistasis-disease-atlas.com). The Epistasis Disease Atlas provides results of eight heritable diseases (see Section Datasets). It enables users to browse, visualize, interpret, and link candidate SNP sets and their interactions within the SSI network to published literature and a variety of external databases and tools (see Methods for details). The Epistasis Disease Atlas can further be queried through an application programming interface (API) where we offer both R and Python packages (Suppl. Materials 2).

## Discussion

The initially high expectations towards genome-wide association studies have thus far not completely been fulfilled due to the missing heritability problem^4, 5^, which suggests that individual genetic variants cannot explain the genetic architecture of complex and polygenic heritable diseases. However, it is likely that the missing heritability is overestimated since epistatic interactions are implicitly ignored in these estimates^37^. While statistical models and computational tools for identifying epistatic interactions have been proposed, the space of possible interactions to be tested is vast and could thus far mostly tackle pairwise interactions. With NeEDL, we overcame this problem and suggest promising higher-order SNP sets. NeEDL outperforms the existing tools LinDen and MACOED, which are limited to pairwise interactions, across all commonly used statistical models and offers new insights into the genetic architecture of complex diseases. We show evidence that candidate SNP sets that are strongly associated with disease phenotypes are in the order of three to seven, are biologically plausible, and can be verified in independent cohorts.

The candidate SNP sets identified by NeEDL represent locally optimal solutions within specific areas of an SSI network and are thus limited by the quality of the priors in this network. While the current version of NeEDL generally outperforms random priors, we see room for further improvement in designing the SSI networks. It is supported by the observation that occasionally, random solutions achieved better scores in our benchmark. For instance, PPI networks such as BioGRID may contain a substantial amount of missing interactions such that a globally optimal SNP set cannot be found by NeEDL. Similarly, existing PPI networks neglect the consequences of alternative splicing on functional interactions^38^, for instance. Furthermore, SNPs that alter codon efficiency during the translation process can reduce or inhibit protein activity^39^, which ultimately can alter a PPI network. More disease-associated variants are found in non-coding compared to coding regions of the DNA, suggesting a major role of gene regulation in human diseases^40^. This motivates further research in SSI networks that consider enhancer-gene interaction and chromatin remodeling. We consider only SNPs that have previously been associated with genes in dbSNP; thus, other associations are neglected. Including gene-regulatory interactions will also pose a significant challenge for interpretability, as the current strategy of GSEA and KEGG enrichment is limited to established interactions between genes and proteins. In summary, the higher the quality of the prior knowledge NeEDL can use to reduce the search space, the more meaningful the results will be.

We should further consider the limitations of the statistical models employed in NeEDL. Recent work by Blumenthal et al.^12^ has demonstrated significant differences in the detection power of various statistical models, concluding that the MLM score offers the best tradeoff between quality and performance, outperforming the quadratic regression model, Bayesian model, and variance model. The *P* -values obtained through the *χ*^2^-test are extremely small (approaching the minimum C++ double value) and preclude further differentiation between candidate SNP sets. Hence, we recommend using the MLM score. Covariates, such as age, co-morbidities, environmental factors, lifestyle factors, and population structure, could negatively impact the discovery of epistatic interactions. While regression models can, in principle, control for such effects (which is also supported by NeEDL), this further increases the computational demand beyond currently available resources. Further algorithmic improvements in mining the SSI network, maybe through graph neural networks, are conceivable and will be a promising research direction^41^. As a potential solution to further speed up the local search algorithm of NeEDL, we considered various quantum algorithms and devices, showing that epistasis detection poses a real-world scenario in which quantum advantage could be achieved and exploited and, in the foreseeable future, allow NeEDL to also handle increasingly complex data sets with millions of SNPs.

Replication of epistasis candidate SNP sets in independent cohorts, as shown here, can provide valuable confirmation of statistical associations and offer a testbed for assessing the clinical potential of epistatic interaction scores similar to polygenic risk scores. To date, multi-SNP epistasis hypotheses are difficult to verify experimentally. In spite of these challenges, future gene editing studies will be needed for their verification and to unravel the functional consequences of epistatic interactions in disease pathophysiology. To optimally support this process, we developed the Epistasis Disease Atlas, a user-friendly and interactive web resource that will allow the biomedical community to extract testable hypotheses directly and work towards an improved understanding of disease mechanisms as the basis for developing targeted therapeutic interventions. We note that the application of NeEDL is wider than just human disease research and see the potential for applying NeEDL in model organisms such as Arabidopsis thaliana or Mus musculus, where gene editing and functional studies are more readily achievable.

In conclusion, NeEDL is an epistasis detection tool to leverage prior biological knowledge for the systematic discovery of higher-order candidate SNP sets and quantum computing. NeEDL thus lays the foundation for charting the complex genetic architecture underlying heritable diseases. NeEDL lifts epistasis detection to a systems-oriented network biology level and shows, as the first ready-to-use real-world application, that seamless integration of quantum computing has the potential to transform computational genomics, highlighting its role in biomedical research.

## Methods

### Implementation details

NeEDL is implemented in C++. It employs the Boost Graph Library version 1.71.0^42^ and iGraph version 0.9.8^43^ for the construction and handling of graphs. We use OpenMP^44^ for the parallelization of the initialization of NeEDL (i.e., read the input data, map the SNPs to genes, construct the SSI network, and get random start seeds) and the local search with simulated annealing. We further included the following external dependencies: CMake v. 2.6 or higher, Doxygen, Catch version 2.11.0, CLI11 version 1.9.0, and Eigen version 3.3.7^45^. We provide an installer script that installs most external dependencies (excluding CMake, Doxygen, and OpenMP). The dockerized version of NeEDL can be directly pulled and executed from: https://hub.docker.com/r/bigdatainbiomedicine/needl.

We conducted computation on the high-performance computing systems provided by the Leibniz Super-computing Center of the Bavarian Academy of Sciences and Humanities (LRZ). Each MACOED task was run using a single core (2.6 GHz nominal frequency) on LRZ’s Large Memory Teramem cluster (HP DL580 shared memory system) since MACOED has extremely high memory requirements (approx. 1.3 terabytes of RAM). For each NeEDL task, we used 28 threads on 14 physical cores (2.6 GHz nominal frequency) on LRZ’s CoolMUC-2 cluster (28-way Haswell-EP nodes).

Since MACOED uses a randomized optimization technique (ant colony optimization), we ran it 100 times on each dataset to minimize the impact of random bias. All obtained SNP sets were used for downstream evaluation. Also, NeEDL includes a randomized subroutine, namely, the seeding of the initial solutions for the local search. To account for this, we ran NeEDL’s local search with multi-start and introduced a global time limit of 12 hours.

The behavior of NeEDL can be influenced through various parameters, including (1) seeding procedure (default: random seeding), (2) maximal size of SNPs in a candidate SNP set (default: 10), (3) maximum iteration of lookups inside one local neighborhood (default: 300), (4) statistical model used for the local search (default: maximum likelihood model), and (5) SNP filters inside the network (e.g., minor allele frequency, maximum marginal association, linkage disequilibrium cutoff, default: None). For further parameters, please check the GitHub repository and the manual of NeEDL.

### Data format converter, data cleaning, and data filtering

We faced challenges in using different genotype and phenotype data formats with NeEDL and other epistasis detection tools due to the lack of available data processing tools for converting these formats to the JSON format required by NeEDL. Furthermore, most epistasis detection tools require formatted, pre-cleaned, and pre-filtered datasets as input. To automate preprocessing, cleaning, filtering, and conversion into a joint machine-readable format, we developed epiJSON which supports VCF, PED/MAP, TPED/TFAM, BED/BIM/FAM, and other formats and follows recommendations laid out by Marees et al.^46^: (1) excluding samples with missing phenotypes; (2) excluding SNPs with more than 20% of missing data across individuals and individuals with more than 20% of missing data across SNPs; (3) checking an individual’s assigned sex using X chromosome data, and removing those with a discrepancy; (4) considering the number of samples within the given dataset to determine a suitable minor allele frequency threshold; (5) removing variants that fail the Hardy-Weinberg test at a threshold recommended for binary traits of 1*e*^*−*10^ in controls and 1*e*^*−*6^ in cases; (6) excluding individuals with heterozygosity that is too high or too low, indicating sample contamination or inbreeding; and (7) excluding SNPs with any missing data across individuals to ensure that the dataset has no missing values. Cleaning processes and filtering steps can be adjusted with parameters following the guideline in the GitHub repository.

### Datasets

Eight datasets were included in our benchmark: Late-Onset Alzheimer’s Disease (LOAD), Bipolar Disorder (BD), Coronary Artery Disease (CAD), Diabetes Type 1 (T1D), Diabetes Type 2 (T2D), Hypertension (HT), Inflammatory Bowel Disease (IBD), and Rheumatoid Arthritis (RA) (see Section Data Availability for links to the databases). The LOAD dataset had controls included. All other datasets had no controls included, and we used the British Cohort 1958 provided by the Wellcome Trust Case Control Consortium (WTCCC) as controls. In (Suppl. Tables 4-12), we show the detailed sample and SNP numbers before and after the epiJSON tool for the datasets discussed in this manuscript. The LOAD dataset was provided by the TGen consortium^19, 47^, the other datasets were provided by the WTCCC consortium^48^ (see Data availability).

A replication study for SNP sets discovered in the disease LOAD was conducted using data from the UK Biobank database (www.ukbiobank.ac.uk)^49^. Individuals were selected using ICD-10 coding^50^ (field 41270), see Suppl. Table 13. We constructed two replication data sets. One containing individuals with mixed ancestry and a sub-data set containing only individuals with British ancestry. Individuals in controls matched the individuals in cases in age (field 34) and gender (field 31) proportionally. Replication was performed on the results of the discovery study using the tools NeEDL, higher-order baseline, second-order baseline, MACOED, and LinDen. The Pearson correlation coefficient was calculated^51^ to determine the correlation of statistical scores between the discovery and replication dataset for the candidate SNP sets for LOAD of the NeEDL.

### Epistasis models

For statistical modeling, we use the framework introduced by Blumenthal *et al*.^12^. Let **G** = (*g*_i,s_) ∈ {0, 1, 2} ^*ℐ× 𝒮*^ be a genotype matrix, where ℐ and 𝒮 denote the sets of all individuals (patients) and SNPs, respectively, and the entry *g*_i,s_ encodes the number of minor alleles of individual *i* at SNP *s*. Moreover, let **y** ∈ ℙ^*ℐ*^ be a phenotype vector, where ℙ is the set of all possible phenotypes (e. g., ℙ = {0, 1} for case-versus-control data). We call a tuple *M* = (*f*_**G**,**y**_, *σ*) an *epistasis model* if *σ* ∈ {*MIN, MAX* } is the model sense and *f*_**G**,**y**_ : 𝒫 (𝒮) → ℝ is an objective function that assigns objective values *f*_**G**,**y**_(*S*) to SNP sets *S* ⊆ 𝒮 which quantify the statistical evidence for the SNPs *s* ∈ *S* being involved in epistatic interaction.

A wide variety of epistasis models have been proposed in the literature, including Bayesian network scores (such as the K2-score^52–57^), variants of multifactor dimensionality reduction^58–62^, variants of polynomial regression^63–65^, the *P* -value of the *χ*^2^-test, which is arguably the most widely used epistasis model^52–57^, or the maximum likelihood model (MLM) introduced by Blumenthal *et al*.^12^.

Equipped with these definitions (and assuming *σ* = *MIN* ; if *σ* = *MAX* we instead have to maximize in (Equation (1)), the problem of detecting SNPs involved in epistatic interaction can, in a first attempt, be formalized as the problem to find an SNP set *S*^*^ that solves the optimization problem

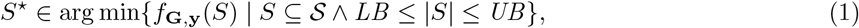

where *LB* ≥2 and *UB* ≥*LB* are user-specified lower and upper bounds on the size of the solution SNP set. The problem with this naïve formulation is that the search space is huge, and solving the optimization problem is hence computationally very expensive. In NeEDL, we mitigate this shortcoming by restricting the search space to SNP sets *S*, which induce a connected subgraph 𝒩 [*S*] in an SSI network 𝒩 = (𝒮, ℰ) where two SNPs *s*_1_, *s*_2_ ∈ 𝒮 are connected by an edge *s*_1_*s*_2_ ∈ ℰ if there exists prior evidence that they might be involved in a biologically meaningful interaction (see the following subsection for details on the construction of 𝒩). That is, NeEDL uses the following computational model:

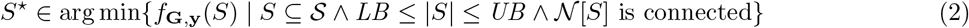

For our benchmark, we use four epistasis models: the *P* -value of the *χ*^2^-test, the K2-score, the MLM score, and the NLL gain of a quadratic regression model in comparison to a linear regression model. The former three models are all based on the penetrance table *T*_**G**,**y**,S_(**g**) = {{*y*_i_| *i*∈ ∐ : *g*_i,s_ = *g*_s_ ∀*s*∈ *S* }} of the scored SNP set *S*. For each possible genotype **g** ∈**{**0, 1, 2}^S^ at the scored SNP set *S*, the cell *T*_**G**,**y**,S_(**g**) of the penetrance table contains the multiset of phenotypes of individuals whose genotype at *S* matches **g**. For all three models, the score *f*_**G**,**y**_(*S*) is small if the phenotypes are unevenly distributed across the cells of *T*_**G**,**y**,S_ (see Blumenthal *et al*.^12^ for details). For the *P* -value of the *χ*^2^-test, the K2-score, and the MLM score, we hence have *σ* = *MIN* and small scores *f*_**G**,**y**_(*S*) indicate that the genotype at *S* is predictive of phenotypic variation.

The NLL gain of quadratic versus linear regression captures another dimension of the concept of epistasis. Here, we first fit two linear or logistic (depending on whether phenotypes are quantitative are categorical) regression models as follows (**G**_*•*,s_ is the column for the SNP *s* in **G** and ⊙ denotes element-wise multiplication):

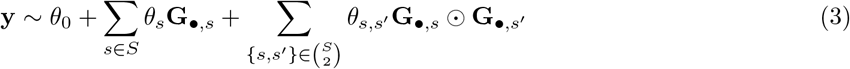

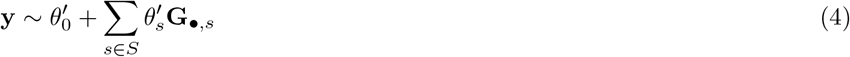

Subsequently, we compute NLLs for the fit models ***θ*** and ***θ′*** and define our score as *f*_**G**,**y**_(*S*) = NLL(***θ′***) NLL(***θ***). For the NLL gain, large scores hence indicate that we can better predict the phenotypes when considering multiplicative interactions between all pairs of SNPs contained in the scored SNP set *S* than when considering only marginal effects (i. e., we have *σ* = *MAX*). Since the NLL gain can result in negative values, we excluded such values from the analysis.

### Construction of the SNP-SNP interaction network

To construct 𝒩= (𝒮, *ℰ*), we require a many-to-many SNP-to-gene mapping *π* ⊆ (𝒮 × 𝒢) (𝒢 is the set of all genes), as well as a gene-gene network 𝒩*′* = (𝒢, *ℰ′*) (Figure 2a). In NeEDL, we obtain *π* from dbSNP^13^ and use the BioGRID^66^ PPI network to construct 𝒩*′*. That is, two genes *g*_1_, *g*_2_∈ 𝒢 are connected by an edge *g*_1_*g*_2_ ∈ ℰ*′* if their encoded proteins are connected by an edge in BioGRID. Note that in BioGRID, two proteins are connected by an edge if and only if a physical interaction between them is supported by at least two publications or two experimental systems (e. g., affinity purification-mass spectrometry and yeast-2-hybrid)^67^. However, NeEDL can be easily extended to use other SNP-to-gene mappings and/or gene-gene networks. For each SNP *s*∈ 𝒮, let *π*[*s*] = {*g*∈ 𝒢 | (*s, g*) ∈*π*} be the image of *s* under the mapping *π*. Equipped with *π* and 𝒩 ′, we construct 𝒩 by connecting to SNPs *s*_1_, *s*_2_∈ 𝒮 by an edge *s*_1_*s*_2_∈ ℰ if and only if at least one of the following conditions holds:

- The SNPs *s*_1_ and *s*_2_ are mapped to at least one common gene, i. e., *π*[*s*_1_] ∩ *π*[*s*_2_] ≠ ∅.
- There is a SNP *s*_3_ and genes *g*_1_, *g*_2_ ∈ *π*[*s*_3_] such that *g*_1_ ∈ *π*[*s*_i_] and *g*_2_ ∈ *π*[*s*_2_].
- The SNPs *s*_1_ and *s*_2_ are mapped to genes that are connected in the gene-gene network 𝒩 ′, i. e., there is an edge *g*_1_*g*_2_ ∈ ℰ′ such that *g*_1_ ∈ *π*[*s*_1_] and *g*_2_ ∈ *π*[*s*_2_].

Note that, with this construction, SNPs that are left unmapped by *π* correspond to isolated nodes in 𝒩 and are hence not contained in any feasible solution of the optimization problem specified in Equation (2). In NeEDL, we hence remove all unmapped SNPs before (heuristically) solving the optimization problem, which further reduces the size of the search space.

### Local search with multi-start and simulated annealing

We use the local search with multi-start and simulated annealing to heuristically solve the optimization problem specified in Equation (2). NeEDL supports all epistasis models benchmarked by Blumenthal *et al*.^12^. Simulated annealing is a general meta-heuristic to escape local optima in local search by accepting deteriorations from local optima with probabilities that decrease as the algorithm runs^68^. For NeEDL, we adapted a simulating annealing algorithm for graph edit distance computation proposed by Riesen *et al*.^69^, using its implementation available in GEDLIB^70, 71^.

Algorithm 1 provides a high-level description of our algorithm. It computes a set 𝒮 ⋆⊆ ℱ_*LB,UB*_ of up to *n* locally optimal SNP sets, where *n* is a hyper-parameter that can be set by the user and ℱ_*LB,UB*_ = {*S* ⊆ 𝒮 | *LB*≤ |*S* | ≤*UB*∧ 𝒩[*S*] is connected is the set of all feasible solutions. Computation of the *n* locally optimal SNP sets is parallelized, which is the main reason for NeEDL’s excellent runtime performance (see “Implementation details” for details).

#### Algorithm 1

Overview of local search and simulated annealing.

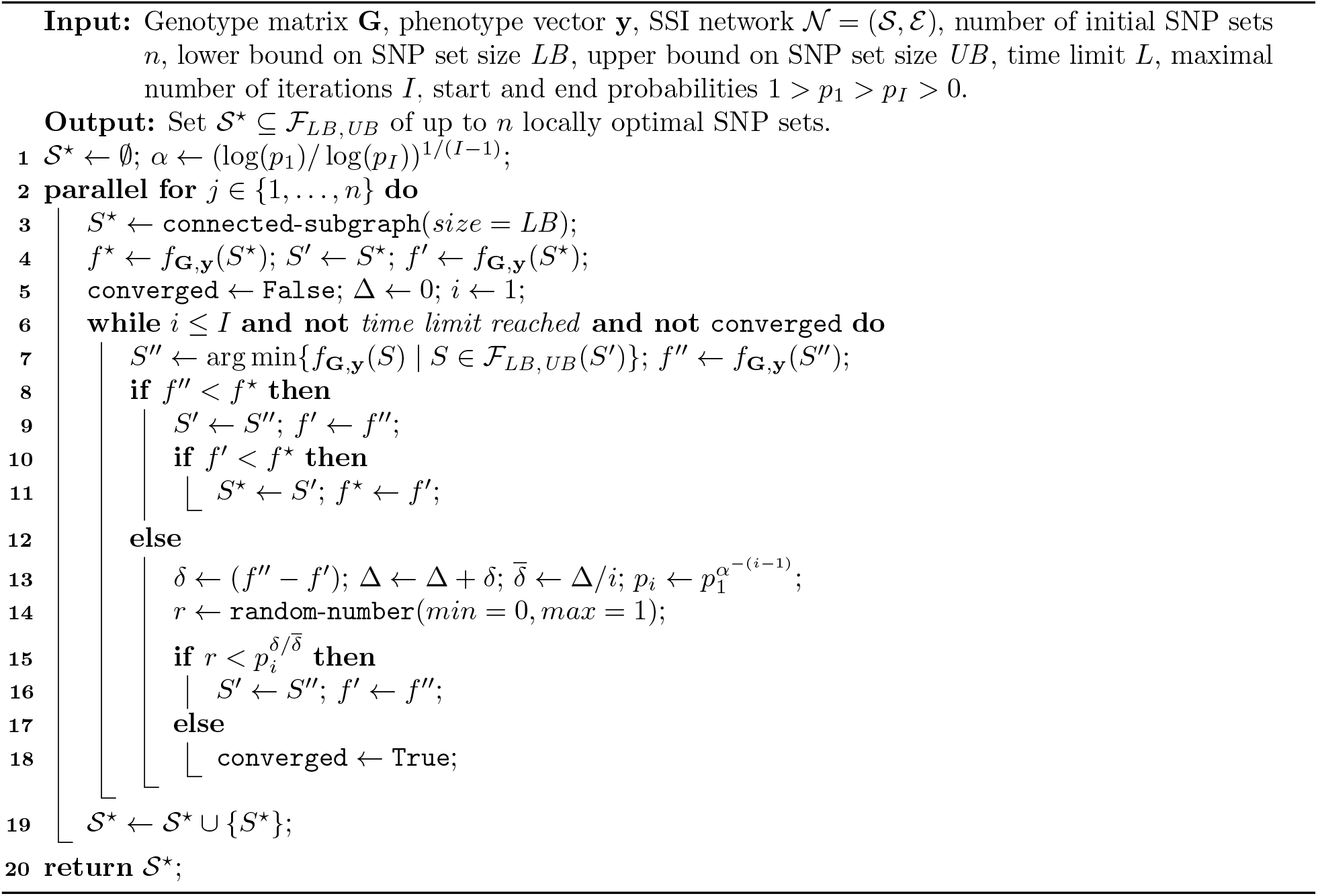

To construct the *j*^th^ locally optimal SNP set, we maintain the best encountered SNP set *S*⋆ (which we initialize as a randomly connected subgraph of size *LB*), as well as the currently processed SNP, set *S′*, together with their objective values *f*⋆ and *f ′*. As long as a given time limit or a maximal number of iterations has not been reached and our descent in the *j*^th^ region of the search space has not converged, we compute a candidate SNP set *S*″ as the best SNP set among the set ℱ_*LB,UB*_ (*S′*) of all locally reachable SNP sets from *S′*. We define ℱ_*LB,UB*_ (*S′*) = {*S* ∈ ℱ | |*S △ S′* | = 1} as the set of all feasible SNP sets that differ from *S′* in exactly one SNP (details on this step are provided below). We then check whether *S*″ improves the currently processed SNP set *S′*. If so, we update *S′* and also *S*⋆ if *S′* is now the best encountered SNP set.

On the other hand, if the objective value *S*″ exceeds the objective value of *S′* by *δ >* 0, we nonetheless accept it as our new currently processed SNP set with probability 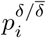, where 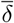 is the mean deterioration from the objective value of the currently processed SNP set up to the current iteration *i*. That is, the larger *δ*, the lower the probability of accepting *S*″as our new currently processed SNP set. The baseline acceptance probability *p*_1_ for the first iteration is a hyper-parameter that can be provided by the user (defaulted to *p*_1_ = 0.8 in NeEDL). As the algorithm runs, it is updated using a suitably defined cooling factor *α* such that, when the maximum number of iterations *I* has been reached, it equals a user-specified end probability *p*_I_ *< p*_1_ (defaulted to *p*_I_ = 0.01 in NeEDL).

To compute *S*″, we exhaustively enumerate the set ℱ_*LB,UB*_ (*S′*) of all locally reachable SNP sets from *S′*. This can be done by constructing modified SNP sets using the following three types of edit operations:

- SNP insertions (only if |*S′* | *< UB*): Insert an SNP *s* ∈ 𝒮\*S′* that is connected to one of the SNPs already contained in *S′*.
- SNP removals (only if |*S′* | *> LB*): Remove a SNP *s* ∈*S′* such that the induced subgraph 𝒩[*S′\* {*s*}] remains connected (i. e., *s* must not be an articulation point).
- SNP substitutions: Substitute a SNP *s*_1_ ∈ *S′* by a SNP *s*_2_ ∈ 𝒮\ *S′* such that the induced subgraph 𝒩[(*S′* \ {*s*_1_}) ∪ {*s*_2_}] remains connected (i. e., if *s*_1_ is an articulation point, *s*_2_ must bridge the connected components resulting from removal of *s*_1_).

### Statistical methods

We generate random SNP sets for empirical estimation of the false-positive rate. For epistasis detection tools other than NeEDL, we sample 1,000 random SNP sets of size two as a baseline, whereas for NeEDL, we estimate the distribution of the number of SNPs in candidate sets based on the findings of our results and then randomly sample 1,000 candidate SNP sets following this size distribution.

We generate two random SSI networks to evaluate the information that can be gained by transforming a PPI network into an SSI network. We either shuffle the labels to obtain a topology-preserving SSI network or shuffle the edges by a probability function to obtain an expected degree-preserving network as discussed in Lazareva *et al*.^72^. Either way, this leads to a network where each SNP has other neighbors in the SSI network that result in the loss of biological information Suppl. Fig. 5a).

All *P* -values obtained by NeEDL and all competitors are corrected with the Benjamini-Hochberg procedure^14^. In addition, we provide a baseline of randomly chosen candidate SNP sets, which SNPs do not need to be connected via the SSI network. The baseline for the competitors contains 1000 randomly sampled sets with the size |*s*| = 2, as all competitors (except PoCos, which return hundreds of SNPs, and since the statistical scores cannot be calculated with such a high number of SNPs, PoCos was excluded) only return candidate SNP sets with the size of two. NeEDL finds candidate SNP sets with variable sizes |*s*| ∈ [2, ∞[.

### Gene set enrichment analysis

Affected genes present in the highest ranks of penetrance SNP combinations for LOAD, IBD, and T1D were subjected to GSEA (http://www.gsea-msigdb.org/gsea/index.jsp, date of last access March 8, 2023) to determine if statistically significant enrichment in specific Gene Ontology (GO) gene sets could be identified^15, 16^. The query gene sets for GSEA are contained in Suppl. File 2.

### Quantum computing

While large, fault-tolerant quantum computers are expected to outperform classical devices, complexity theory suggests that quantum computing may not be able to solve combinatorial optimization problems in polynomial time, similar to classical computing. Nevertheless, Grover’s algorithm^73^ can provide a quadratic speedup in terms of query complexity for quantum circuit-based quantum hardware (Suppl. Materials 4), and quantum annealers (Suppl. Materials 4) can provide the same speedup^74^, under ideal, noiseless conditions. For particular instances of NP-hard optimization problems, super-polynomial speedups are possible^75^, and certain problems can achieve even larger speedups by exploiting their specific structural features^76^. Currently, only prototypes of quantum hardware are available, and heuristics must be run on them, with satisfactory results depending on the problem^77^.

We propose a novel quantum-enhanced method to address the initial seeding problem in NeEDL. Specifically, instead of selecting the initial seed randomly, our method employs an optimization technique to identify the most promising candidates. The underlying idea is that a better initial seed can significantly improve the quality of local search and reduce the time required to obtain high-quality solutions, assuming the optimization technique adds negligible overhead. To achieve this, we utilize a variant of the Max-Clique problem to analyze the matrix of pairwise SNP correlations and identify the candidate set of SNPs that maximizes the sum of pairwise correlations. This problem is formulated as a Quadratic Unconstrained Binary Optimization (QUBO) whose objective function to minimize is defined as a quadratic polynomial over binary variables and consists of a linear combination of the solution cost and the associated constraints. By adjusting the weights of the linear combination, we can enforce or relax different constraints, such as the size of the candidate set. We formulate the QUBO problem as follows:

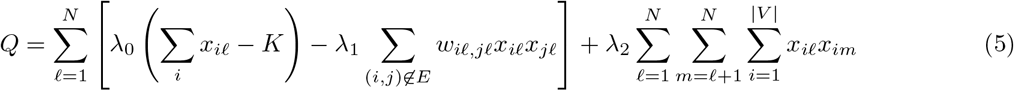

The formulation returns, saved in the matrix of binary variables *x, N* candidate sets of SNPs, each consisting of *K* SNPs. The parameter *λ*_0_ penalizes candidate sets that are not of size *K, λ*_1_ rewards sets with a higher sum of pairwise interactions *w*, and *λ*_2_ penalizes solutions with similar sets. For additional details, see Suppl. Materials 5. We solve the optimization problem by applying quantum annealing on a D-Wave quantum annealer, QAOA (quantum approximate optimization algorithm) on IBM Perth and IBM Lagos quantum processing units, and thermal annealing on classical hardware as a classical baseline. These algorithms are heuristics, and their potential speedup must be determined empirically on a case-by-case basis. The functioning of these quantum algorithms is detailed in Suppl. Materials 6.

To address the limitations in quantum hardware resources, we utilize the community-detection method Leiden^78^ to identify densely connected sections of arbitrary size in the SNPs network. This approach allows us to divide the problem into smaller sub-instances, enabling efficient computation on any quantum hardware. Specifically, quantum annealing algorithms can handle problem sizes on the order of 100 SNPs, while IBM quantum processing units can manage up to 10 SNPs. Our quantum software module called *quepistasis*, implements the seeding procedure using the optimization technique. It is responsible for creating the QUBO formulation based on the pairwise correlation of SNPs and some configuration, interfacing with the platforms of D-Wave and IBM (and also supporting quantum hardware available on Microsoft Azure), and returning the set of SNPs. Its structure is shown in Suppl. Materials 7.

We test NeEDL with the quantum computing seeding procedure over three different datasets: BD, RA, and T2D. The purpose of our experiments is to verify how much quantum computing helps in finding better seeds and thus speeds up the local search process. To accommodate the limited amount of resources (namely the number of qubits and the number of gates) of the quantum computer, we subsample each of the three datasets into three different subsets, having 100, 500, and 1000 SNPs. Minimizing the quantum resources is essential for keeping the *fidelity*, which is the measure of the accuracy of the actual quantum operation compared to the ideal one, above a certain threshold, guaranteeing reliable results. The timing information in Suppl. Fig. 8 is obtained from the logs generated by NeEDL. For the quantum procedure run on IBM devices, such timing is spoiled by the queue we must wait to execute each task, and such time is normalized. The scalability discussion in Figure 5 is obtained by linear interpolation of the seeding time using each technique for different sample sizes of each dataset. We show the experimental setup and provide some comments about the results obtained in Suppl. Materials 8.

## Supporting information

Supplementary Data is available at medRxiv online.

## Data Availability

The LOAD data set is restricted and can be found at https://www.tgen.org. The users need to apply for the data set.
The BD (EGAD00000000003), CAD (EGAD00000000004), T1D (EGAD00000000008), T2D (EGAD00000000009), HT (EGAD00000000006), IBD (EGAD00000000005), RA (EGAD00000000007), and the British 1958 British Birth Cohort (EGAD00000000001) data sets are restricted and can be found at https://www.sanger.ac.uk/legal/DAA/MasterController https://edam.sanger.ac.uk/#/. The user needs to apply for the data sets.
In the future, we plan to include the following datasets in the Epistasis Disease Atlas: AS (EGAD00000000010), ATD (EGAD00000000011), MS (EGAD00000000012), BRCA (EGAD00000000013), TB (EGAD00000000016), UC (EGAD00000000025), PD (EGAD00000000057), AK (EGAD00010000150), SP (EGAD00010000262), IS (EGAD00010000264), CD (EGAD00010000246); these data sets are restricted and can be found at https://www.sanger.ac.uk/legal/DAA/MasterController.
The user needs to apply for the data sets.
The datasets for replication are restricted and can be found at UK Biobank database (www.ukbiobank.ac.uk), project IDs 32683 and 54273.
The results of NeEDL stored in the Epistasis Disease Atlas are freely available under the CC BY-NC 4.0 license.
The source code of NeEDL, the quantum computing module, and the R Shiny App is freely available under the GPLv3 license at GitHub: https://github.com/biomedbigdata/NeEDL.
All features of NeEDL are dockerized and available at Dockerhub: https://hub.docker.com/r/bigdatainbiomedicine/needl

https://epistasis-disease-atlas.com/home

## Data availability

The LOAD data set is restricted and can be found at https://www.tgen.org. The users need to apply for the data set.

The BD (EGAD00000000003), CAD (EGAD00000000004), T1D (EGAD00000000008), T2D (EGAD00000000009), HT (EGAD00000000006), IBD (EGAD00000000005), RA (EGAD00000000007), and the British 1958 British Birth Cohort (EGAD00000000001) data sets are restricted and can be found at https://www.sanger.ac.uk/legal/DAA/MasterController https://edam.sanger.ac.uk//. The user needs to apply for the data sets.

In the future, we plan to include the following datasets in the Epistasis Disease Atlas: AS (EGAD00000000010), ATD (EGAD00000000011), MS (EGAD00000000012), BRCA (EGAD00000000013), TB (EGAD00000000016), UC (EGAD00000000025), PD (EGAD00000000057), AK (EGAD00010000150), SP (EGAD00010000262), IS (EGAD00010000264), CD (EGAD00010000246); these data sets are restricted and can be found at https://www.sanger.ac.uk/legal/DAA/MasterController. The user needs to apply for the data sets. The datasets for replication are restricted can be found at UK Biobank database (www.ukbiobank.ac.uk), project IDs 32683 and 54273.

The results of NeEDL stored in the Epistasis Disease Atlas are freely available under the CC BY-NC 4.0 license.

## Code availability

The source code of NeEDL, the quantum computing module, and the R Shiny App is freely available under the GPLv3 license at GitHub: https://github.com/biomedbigdata/NeEDL.

All features of NeEDL are dockerized and available at Dockerhub: https://hub.docker.com/r/bigdatainbiomedicine/needl

## Acknowledgments

We want to thank Christina Trummer for the design of the NeEDL logo and the Epistasis Disease Atlas logo, as well as the design of the webpage. We further want to thank Simona Dedurkeviciute for her advice on webpage design and architecture. Images were created with https://www.biorender.com. Parts of the images were designed using resources from Flaticon.com under a paid license. ChatGPT version 4, under a paid license, was used to rephrase parts of this manuscript. The database diagrams were created with the support of the LucidApp, which gratefully provided us with a free premium account. We would like to thank Matt Huentelman from The Translational Genomics Research Institute in Phoenix, Arizona, for sharing the TGen II cohort data (LOAD).

This study makes use of data generated by the Wellcome TrustCase-ControlConsortium. A full list of the investigators who contributed to the generation of the data is available from www.wtccc.org.uk. Funding for the project was provided by the Wellcome Trust under awards 076113, 085475, and 090355. The authors gratefully acknowledge the Leibniz Supercomputing Centre for funding this project by providing computing time and support on its Linux Cluster. The authors gratefully acknowledge the Gauss Centre for Supercomputing e.V. (www.gauss-centre.eu) for funding this project by providing computing time on the GCS Supercomputer SuperMUC-NG at Leibniz Supercomputing Centre (www.lrz.de). We acknowledge the CINECA award under the ISCRA initiative, for the availability of high-performance computing resources and support (QPU D-Wave). Access to the IBM Quantum Services was obtained through the IBM Quantum Hub at CERN. The views expressed are those of the authors and do not reflect the official policy or position of IBM, the IBM Q team. We acknowledge support from Microsoft’s Azure Quantum for providing credits and access to the quantum hardware used in this paper.

## Funding

This work was supported in part by the Technical University Munich – Institute for Advanced Study, funded by the German Excellence Initiative. This work was supported in part by the Intramural Research Programs (IRPs) of the National Institute of Diabetes and Digestive and Kidney Diseases (NIDDK). MG is supported by CERN through the CERN Quantum Technology Initiative. AMA is supported by Foundation for Polish Science (FNP), IRAP project ICTQT, contract no. 2018/MAB/5, co-financed by EU Smart Growth Operational Programme. This work was supported by the German Federal Ministry of Education and Research (BMBF) within the framework of the *e:Med* research and funding concept (*grants 01ZX1908A / 01ZX2208A* and *grants 01ZX1910D / 01ZX2210D*). This project has received funding from the European Union’s Horizon 2020 research and innovation programme under grant agreement No 777111. This publication reflects only the authors’ view and the European Commission is not responsible for any use that may be made of the information it contains. Funded by the Deutsche Forschungsgemeinschaft (DFG, German Research Foundation) – 422216132. The research of LI and LSCHU is partially funded by the Bavarian State Ministry of Science and the Arts as part of the Munich Quantum Valley.

## Author contributions

MHO designed and developed most parts of the software architectural concepts, database design, and features of epiJSON, NeEDL, calculate scores, the quantum-computing module, the Epistasis Disease Atlas, and the R Shiny App of the Epistasis Disease Atlas. MHO implemented the initial idea, conducted several analyses, wrote the manuscript, and held the role of the project manager and project leader. JP implemented large parts of NeEDL, benchmarked NeEDL against the competitors, and implemented the concept of the real-time calculation of the scores for the Epistasis Disease Atlas. MI implemented the quantum computing module of NeEDL and conducted several analyses on various quantum-computing hardware. SB conducted literature research, applied for most of the data sets, and implemented the R Shiny App of the Epistasis Disease Atlas. MHO and SB produced the overview video on the EDA. AF, AGP, and KB conducted the replication studies of the potential candidate SNP sets in independent cohorts. AM and CH helped with the technical conceptualization and implemented the backend (database, API, and file server) of the Epistasis Disease Atlas. MHA implemented and designed the front end of the Epistasis Disease Atlas. CH implemented the automatic pipeline to integrate new data sets into the database of the Epistasis Disease Atlas. NT implemented the features of the Integrated Genome Browser. KA implemented the R and Python packages of the Epistasis Disease Atlas. MP and MW conceptualized and executed the big memory runs and long-time runs of NeEDL and MACOED on the LRZ supercluster. ES implemented the epiJSON tool and the linkage-disequilibrium options of NeEDL. ES further reformatted the datasets of the eight discussed diseases into NeEDL readable format. PW, HF, and SK helped with analyses. MVH implemented the co-variate and ancestry options. IL tested the software of NeEDL and the Epistasis Disease Atlas for bugs. LSCHW and LHAF conducted further research on epistatic SNPs in regulatory elements. GG and AA processed the raw Illumina data for the additional ten datasets to NeEDL readable format that are about to be integrated into the Epistasis Disease Atlas but not further discussed in this manuscript. LHAC and OT conducted further research on epistatic SNPs in alternative splicing site regions. NW designed the architecture of the Figures and revised the manuscript. SGL and GC conducted further research on epistatic SNPs in structural biology and binding sites. MC conducted further research on epistatic SNPs that changes translational speed. JJ, HKL, PF, and LHEN conducted biological plausibility studies and drafted parts of the manuscript. AMA performed the theoretical analysis for the quantum computing module of NeEDL. FM, HCG, and KVS provided expertise in epistasis detection throughout the project, suggested algorithmic changes, and revised the manuscript. LSCHU, LI, and MG provided quantum-computing expertise and quantum-computing hardware and helped calibrate the systems. LW and DE provided expertise on the math for the co-variant and mixed-ancestry options of NeEDL. ADP, AMA, and MG supervised the quantum-computing part of NeEDL. PF and LHEN supervised the biomedical part of the project. TK, JB, ML, and DB jointly supervised the project. JB and TK had the initial idea for network-enhanced higher-level epistasis detection. DB had crucial ideas regarding algorithmic optimization solutions. JB and DB first suggested the use of quantum computing. DB implemented the statistical models and, together with MHO, wrote most parts of the manuscript. This project took more than four and a half years to implement, execute, and evaluate - which is the reason for eight first authors and seven senior authors. All authors revised the manuscript and gave their approval.

## Competing interests

During the course of the project, HCG became a full-time employee of Novo Nordisk Ltd.

