## Supplementary Data is available at medRxiv online. for "Network medicine-based epistasis detection in complex diseases: ready for quantum computing"

[1]Chair of Experimental Bioinformatics, TUM School of Life Sciences, Technical University of Munich, Germany
[2]Institute for Advanced Study (Lichtenbergstrasse 2 a, D-85748 Garching, Germany), Technical University of Munich, Germany
[3]National Institute of Diabetes, Digestive, and Kidney Diseases, Bethesda, MD 20892, United States of America
[4]Institute for Computational Systems Biology, University of Hamburg, Germany
[5]Dipartimento di Informatica, Universit'a di Verona, Strada le Grazie 15 - 34137, Verona, Italy
[6]Department of Health Technology, Section for Bioinformatics, Technical University of Denmark, DTU, 2800 Kgs. Lyngby, Denmark
[7]Copenhagen Prospective Studies on Asthma in Childhood (COPSAC), Herlev and Gentofte Hospital, University of Copenhagen, Copenhagen, Denmark
[8]Computational Mass Spectrometry, Technical University of Munich, Freising, Germany
[9]Division Data Science in Biomedicine, Peter L. Reichertz Institute for Medical Informatics, Technische Universität Braunschweig and Hannover Medical School, Rebenring 56, 38106 Braunschweig, Germany
[10]Braunschweig Integrated Centre of Systems Biology (BRICS), Technische Universität Braunschweig, Rebenring 56, 38106 Braunschweig, Braunschweig, Germany
[11]School of Biological Sciences and Technology, Chonnam National University, Gwangju, Korea
[12]Department of Chemistry, Gwangju Institute of Science and Technology, Gwangju, Korea
[13]Department of Biology, Brigham Young University, Provo, UT, USA
[14]International Centre for Theory of Quantum Technologies, University of Gdańsk, 80-309 Gdańsk, Poland
[15]BIO3 - Systems Genetics; GIGA-R Medical Genomics, University of Liège, Liège, Belgium
[16]BIO3 - Systems Medicine; Department of Human Genetics, KU Leuven, Leuven, Belgium
[17]Leibniz Supercomputing Centre of the Bavarian Academy of Sciences and Humanities (LRZ), Garching b. München, Germany
[18]RIKEN Center for Advanced Intelligence Project, Tokyo, Japan
[19]Institute of Clinical Molecular Biology, Christian Albrechts University of Kiel, Kiel, Germany.
[20]European Organization for Nuclear Research (CERN), Geneva 1211, Switzerland
[21]Departments of Oncology & Medicine, Georgetown University, Washington, DC, USA
[22]Computational BioMedicine Lab, University of Southern Denmark, Denmark
[23]Department Artificial Intelligence in Biomedical Engineering (AIBE), Friedrich-Alexander University Erlangen-Nürnberg (FAU), Erlangen, Germany
[●]The authors wish to be known that, in their opinion, the first eight authors should be considered as shared first authors
[†]The authors wish to be known that, in their opinion, the last seven authors should be considered as shared last authors
[*]Contact: ``, ``

### Abstract

Most heritable diseases are polygenic. To comprehend the underlying genetic architecture, it is crucial to discover the clinically relevant epistatic interactions (EIs) between genomic single nucleotide polymorphisms (SNPs)[1–3]. Existing statistical computational methods for EI detection are mostly limited to pairs of SNPs due to the combinatorial explosion of higher-order EIs. With NeEDL (**ne**twork-based **e**pistasis **d**etection via **l**ocal search), we leverage network medicine to inform the selection of EIs that are an order of magnitude more statistically significant compared to existing tools and consist, on average, of five SNPs. We further show that this computationally demanding task can be substantially accelerated once quantum computing hardware becomes available. We apply NeEDL to eight different diseases and discover genes (affected by EIs of SNPs) that are partly known to affect the disease, additionally, these results are reproducible across independent cohorts. EIs for these eight diseases can be interactively explored in the Epistasis Disease Atlas (`https://epistasis-disease-atlas.com`). In summary, NeEDL is the first application that demonstrates the potential of seamlessly integrated quantum computing techniques to accelerate biomedical research. Our network medicine approach detects higher-order EIs with unprecedented statistical and biological evidence, yielding unique insights into polygenic diseases and providing a basis for the development of improved risk scores and combination therapies.

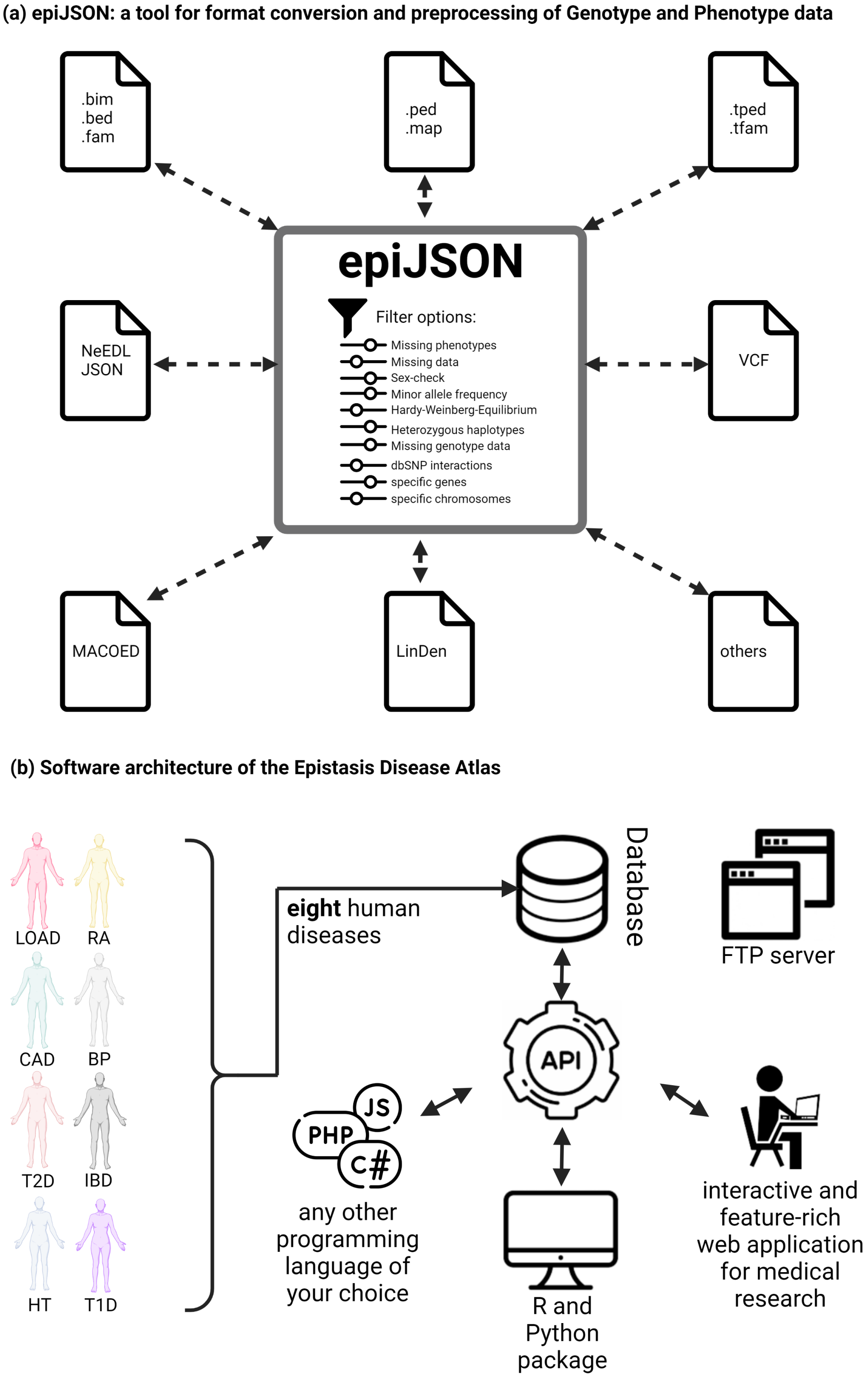


Figure 1: (a) NeEDL supports with its built-in epiJSON tool various widely used GWAS data formats (e.g., the PLINK formats[4]) and provides several optional preprocessing routines[5]. (b) The software architecture of the Epistasis Disease Atlas. The web-based Epistasis Disease Atlas (`https://epistasis-disease-atlas.com`) contains the most promising SNP sets for eight diseases and can be accessed via various access routes.

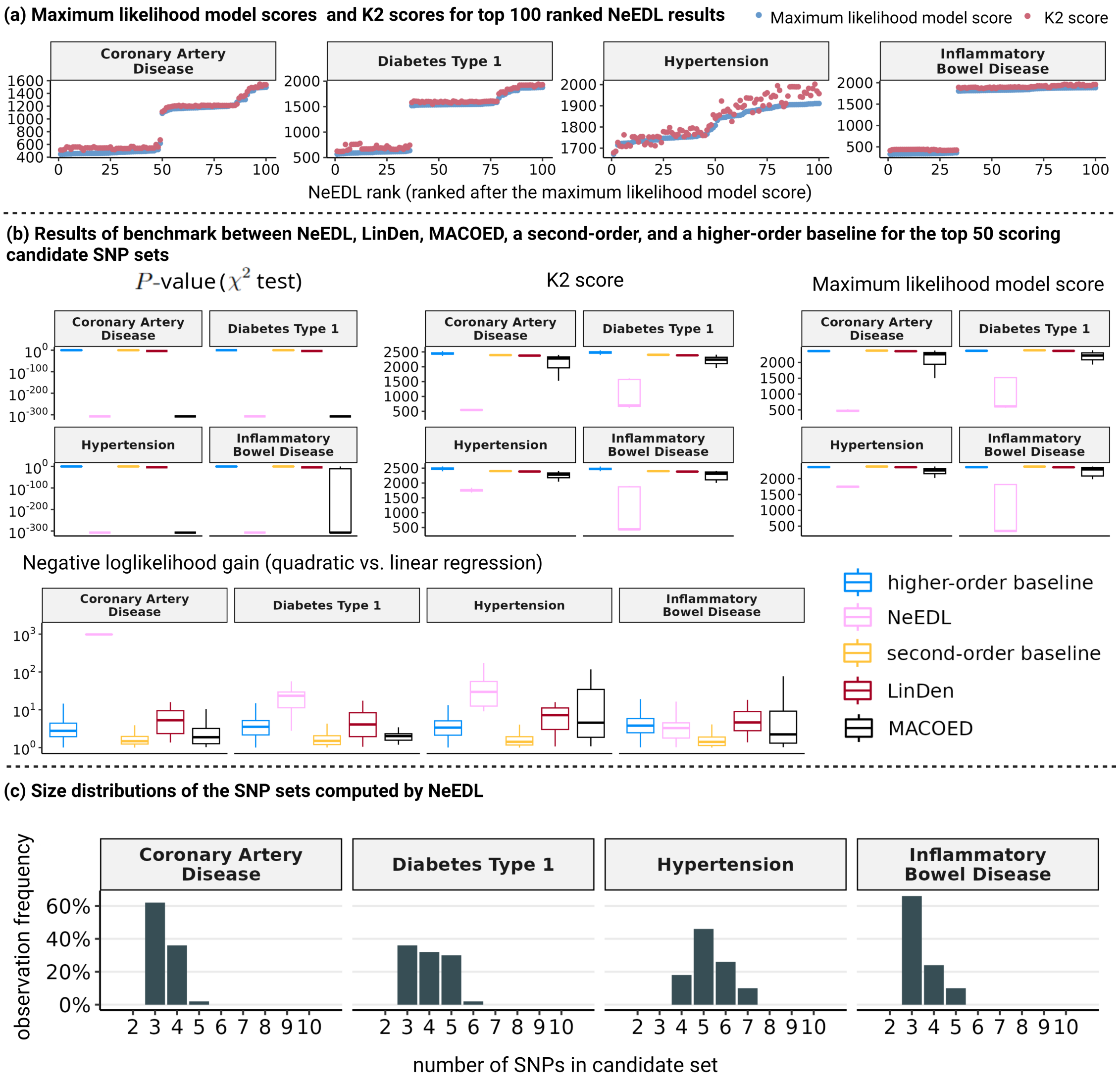


Figure 2: Quantitative evaluation of the SNP sets computed by NeEDL. (a) Visualization of the maximum likelihood model score and the K2 score of the top 100 candidate SNP sets ranked by NeEDL. (b) A benchmark study between NeEDL, LinDen, MACOED, a second-order baseline, and a higher-order baseline shows that NeEDL outperforms in statistical significance existing epistasis detection tools with respect to four different evaluation metrics. (c) Analyzing the number of SNPs included in NeEDL's output, SNP sets reveal that the most promising SNP sets are typical of sizes between three to seven. (d) Comparing maximum likelihood model scores of NeEDL results against those obtained using randomized networks demonstrates that the use of the SSI network indeed leads to the discovery of more promising SNP sets.

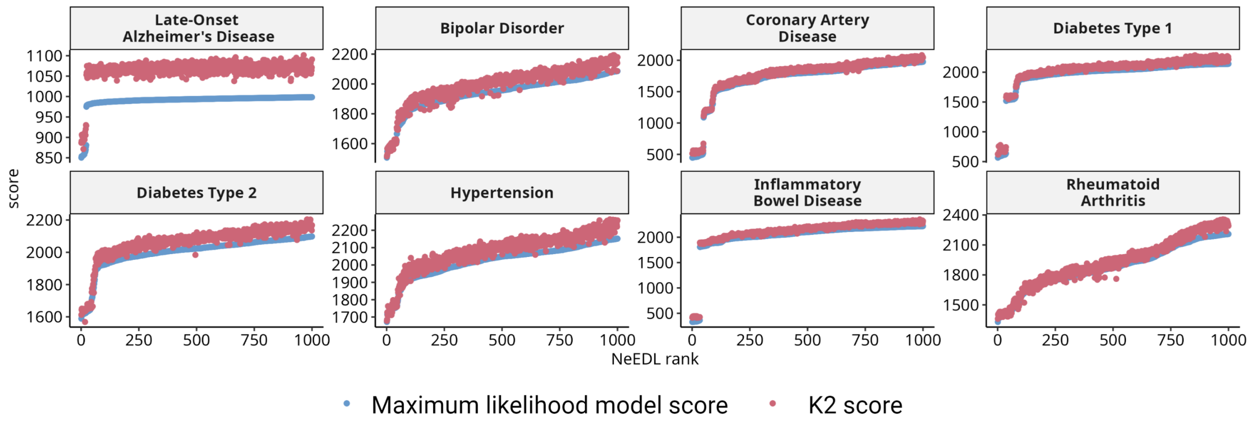


Figure 3: Visualization of the Maximum likelihood model score and the K2 score of the top 1000 candidate SNP sets ranked by NeEDL.

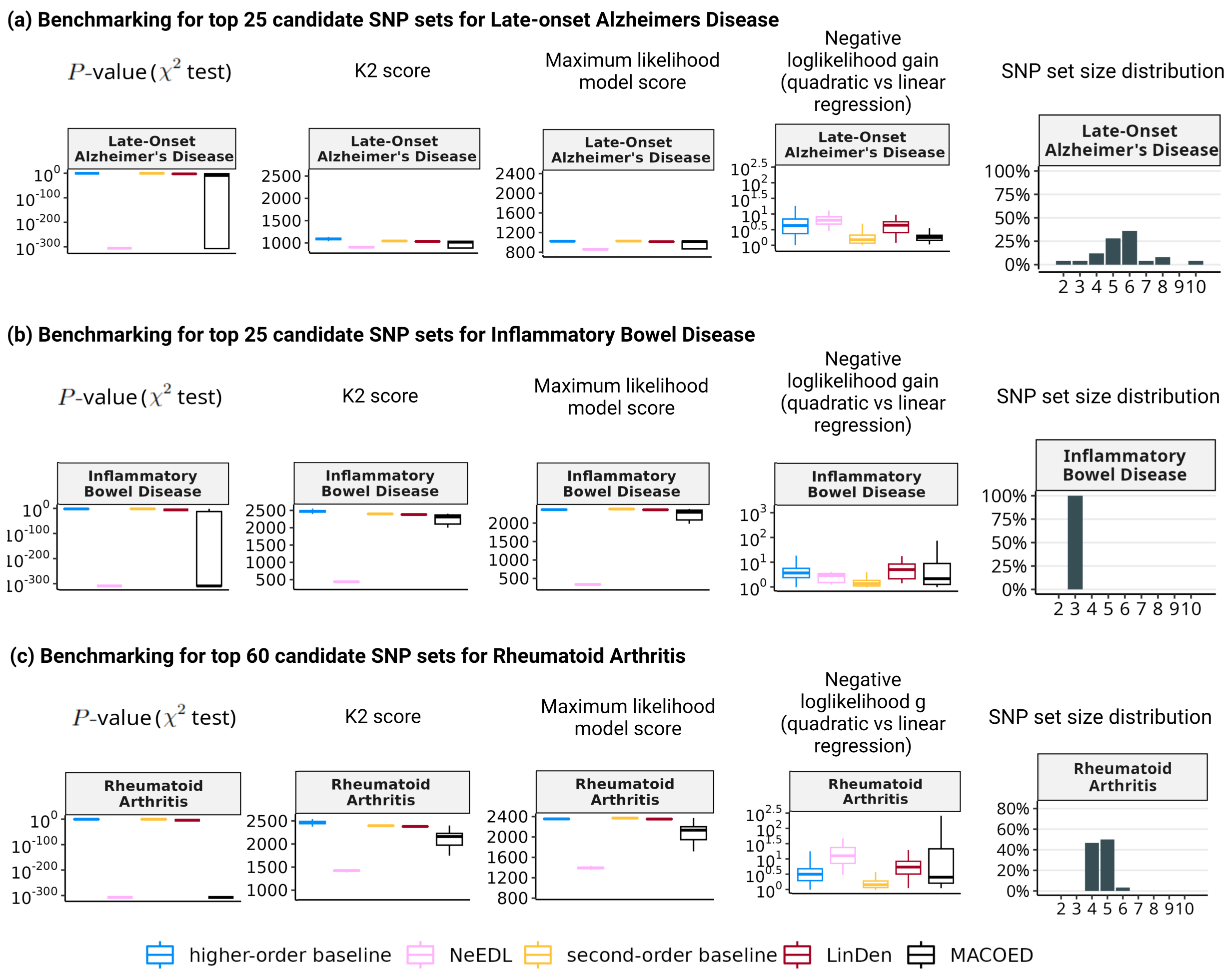


Figure 4: Benchmarking study for other cutoffs for Late-onset Alzheimer's Disease, Inflammatory Bowel Disease, and Rheumatoid Arthritis due to differences of the phase transition gap. (a) A benchmark study for Late-onset Alzheimer's disease between the top 25 ranking candidate SNP sets of NeEDL, LinDen, MACOED, a second-order baseline, and a higher-order baseline shows that NeEDL outperforms LinDen overall metrics and MACOED with respect to the $P$-values and the NLL gain. NeEDL performs similarly or slightly better than MACOED in terms of the K2 score and the MLM score. The resulting SNP set size distribution is shown. (b) A benchmark study for Inflammatory Bowel Disease between the top 25 ranking candidate SNP sets of NeEDL, LinDen, MACOED, a second-order baseline, and a higher-order baseline shows that NeEDL outperforms LinDen in all but the NLL gain metric where it performs similarly. MACOED is outperformed with respect to $P$-values, K2 score, and MLM score but performs similarly in the NLL gain score. The resulting SNP set size distribution is shown. (c) A benchmark study for Rheumatoid Arthritis between the top 60 ranking candidate SNP sets of NeEDL, LinDen, MACOED, a second-order baseline, and a higher-order baseline shows that NeEDL outperforms LinDen in all metrics. MACOED is outperformed in the K2 score, the MLM score, and the NLL gain score but behaves similarly with respect to the $P$-values. The resulting SNP set size distribution is shown.

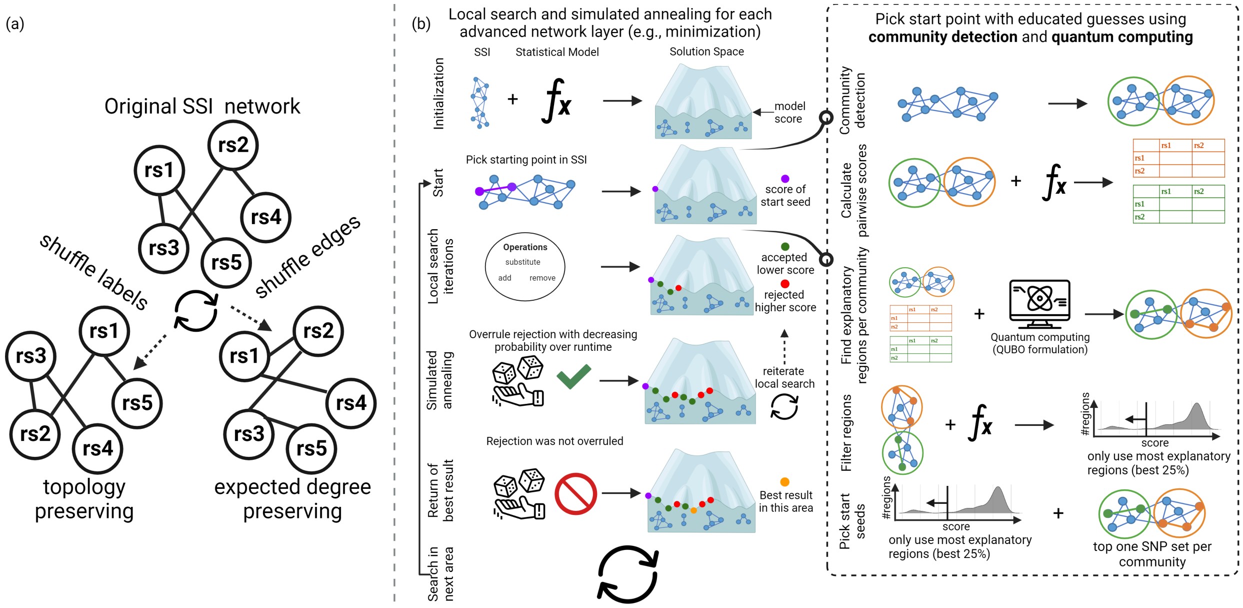


Figure 5: **Overview of a) generation of random background models and b) the heuristics of local search and simulated annealing with quantum computing seeding.** a) We generate two random background models to evaluate the information that can be gained by transforming a PPI network into an SSI network. We either shuffle the labels to obtain a topology-preserving network or shuffle the edges to obtain an expected degree-preserving network. b) After initialization using the SSI network and a statistical model, we let the algorithm pick starting seeds (i.e., connected sets of SNPs either by random or by quantum computing). If we use quantum computing, we first identify densely connected communities in the network and calculate all pairwise scores of the community in a matrix. The matrix is then used in a QUBO formulation that can identify already promising regions in a community. We filter for the 25% most promising regions and use them as starting seeds. Next, we add, substitute, or remove SNPs from the set and the direct neighborhood in the SSI network until we cannot alleviate the score and rejects any neighboring solutions. To avoid being trapped in local minima, we employ simulated annealing that allows us to overrule a rejection with a decreasing probability over the time that is already being used in investigating this area. If we overrule the rejection, we reiterate the local search until the rejection was not overruled anymore. Lastly, we report the best result in this area and start to search for the most promising solution in the next area of the SSI.

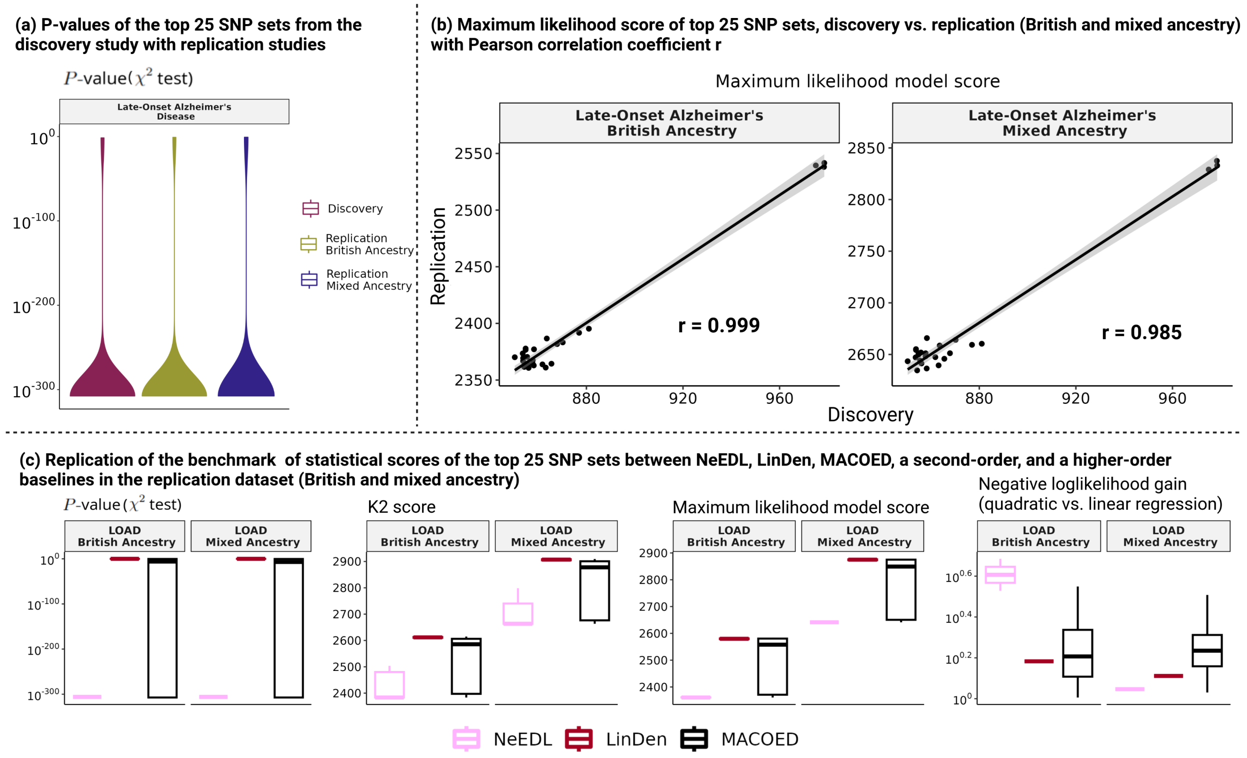


Figure 6: Replication studies in two independent data sets from UK Biobank with British and mixed ancestry. (a) $P$-values across discovery and the two replication studies. (b) Correlation of the MLM score between the discovery study and the two replication studies. (c) Replication of the benchmarking between NeEDL, LinDen, MACOED, a second-order baseline, and a higher-order baseline in two replication data sets.

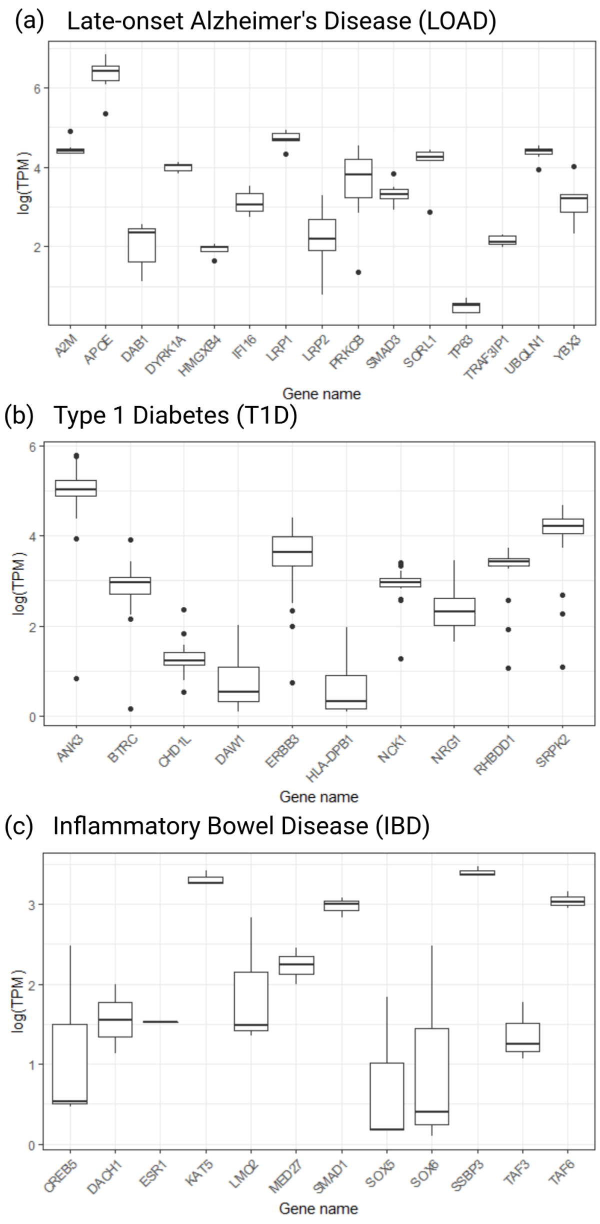


Figure 7: **Tissue-specific transcript per million (TPM) expression of genes enriched in critical pathways involved in disease development.** (a) We included TPM values of the genes APOE, SORL1, SMAD3, LRP2, YBX3, TP63, PRKCB, DAB1, LRP1, IFI16, UBQLN1, A2M, DYRK1A, HMGXB4, and TRAF3IP1 of the tissues (Allocortex, Ammon's Horn, Brainstem, Cerebellum, Diencephalic nuclei, Entorhirnal region, Hippocampus, Insular cortex, Neocortex, Temporal neocortex[6–8]) that were reported as affected in the literature in LOAD from the Human Protein Atlas[9]. (b) We included TPM values of the genes NCK1, ANK3, DAW1, BTRC, CHD1L, NRG1, ERBB3, HLA-DPB1, RHBDD1, SPRK1 of the tissues (Cerebellum, Endothelial cells, Heart, Kidney, Myelin, Pancreatic beta cells, Putamen, Retinal cells, Right parahippocampal gyrus, Thalamus[10–16]) that were reported as affected in the literature in T1D from the Human Protein Atlas. (c) We included TPM values of the genes TAF6, SMAD1, TAF3, KAT5, ESR1, MED27, SSBP3, LMO2, PASD1, DACH1, SOX5, SOX6, and CREB5 from tissues (Blood vessels, Colonic mucosa, Intestinal mucosa, Muscularis external, Serosa, Submucosa[17–19]) that were reported as affected in the literature in IBD from the Human Protein Atlas.

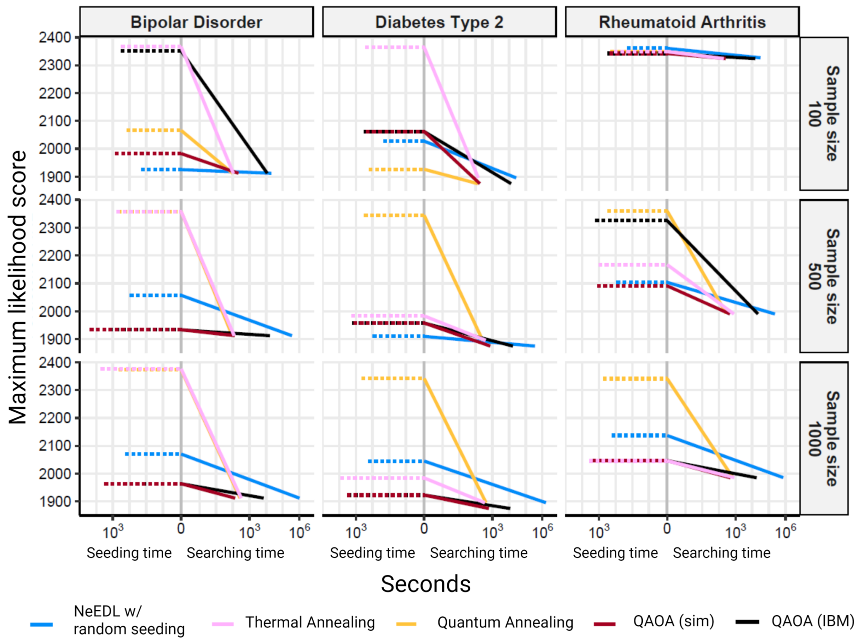


Figure 8: Experiments on the quantum computers. The sample size refers to the number of SNPs. Comparison of the timing performance between Linear NeEDL and Quantum-computing enabled NeEDL. The negative time indicates the duration seeding process, and the positive time is the searching process. The seeding with quantum computing produces fewer but better seeds, speeding up the local search phase.

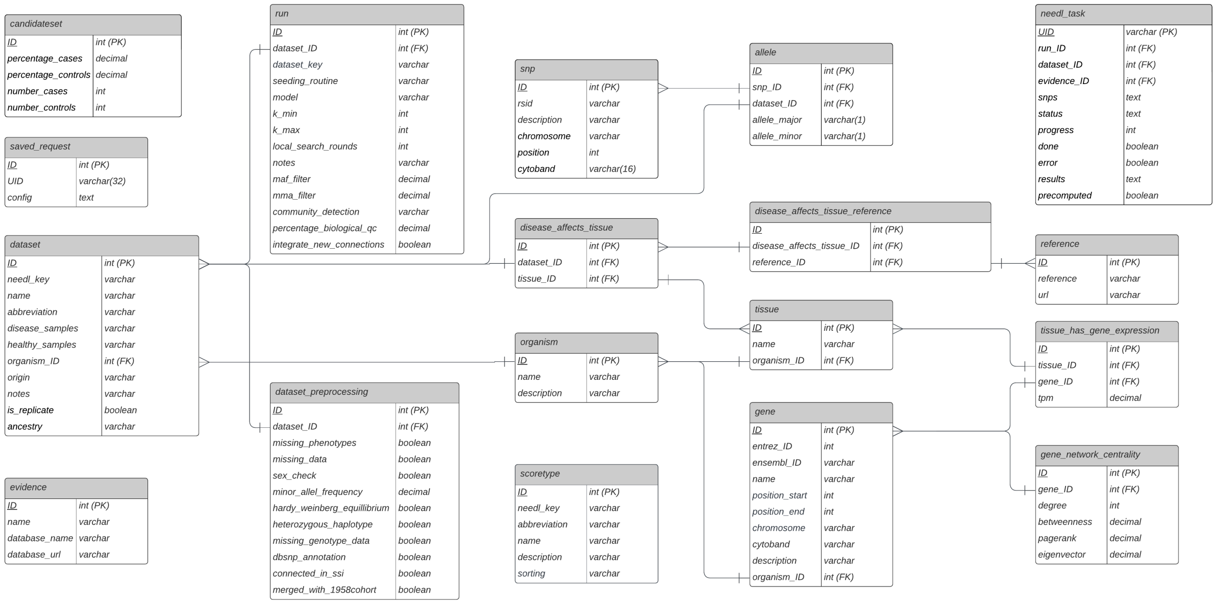


Figure 9: An UML diagram of the Epistasis Disease Atlas SQL database. The layout roughly reflects the origin of information saved in each table. Left-oriented are tables with information specific to the project, whereas, to the right, generally available information to annotate given data is shown.

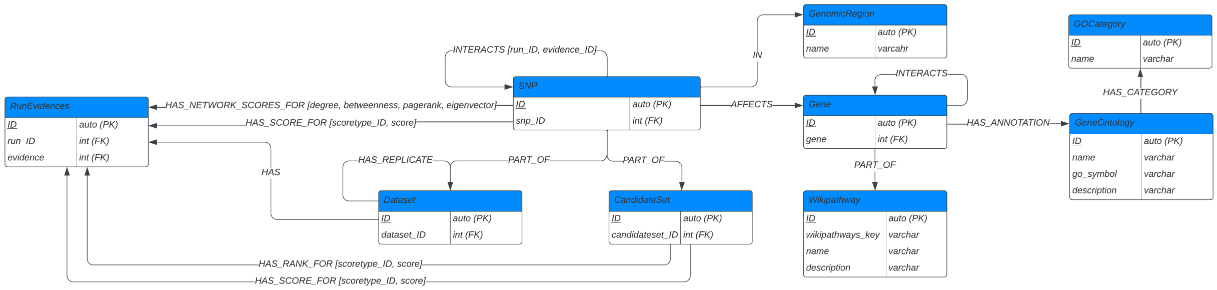


Figure 10: An UML diagram of the Epistasis Disease Atlas Neo4j database. Nodes (entities) are represented as tables and edges (relationships) as connections between tables. The database design is fully focused on executing necessary queries time-efficiently.

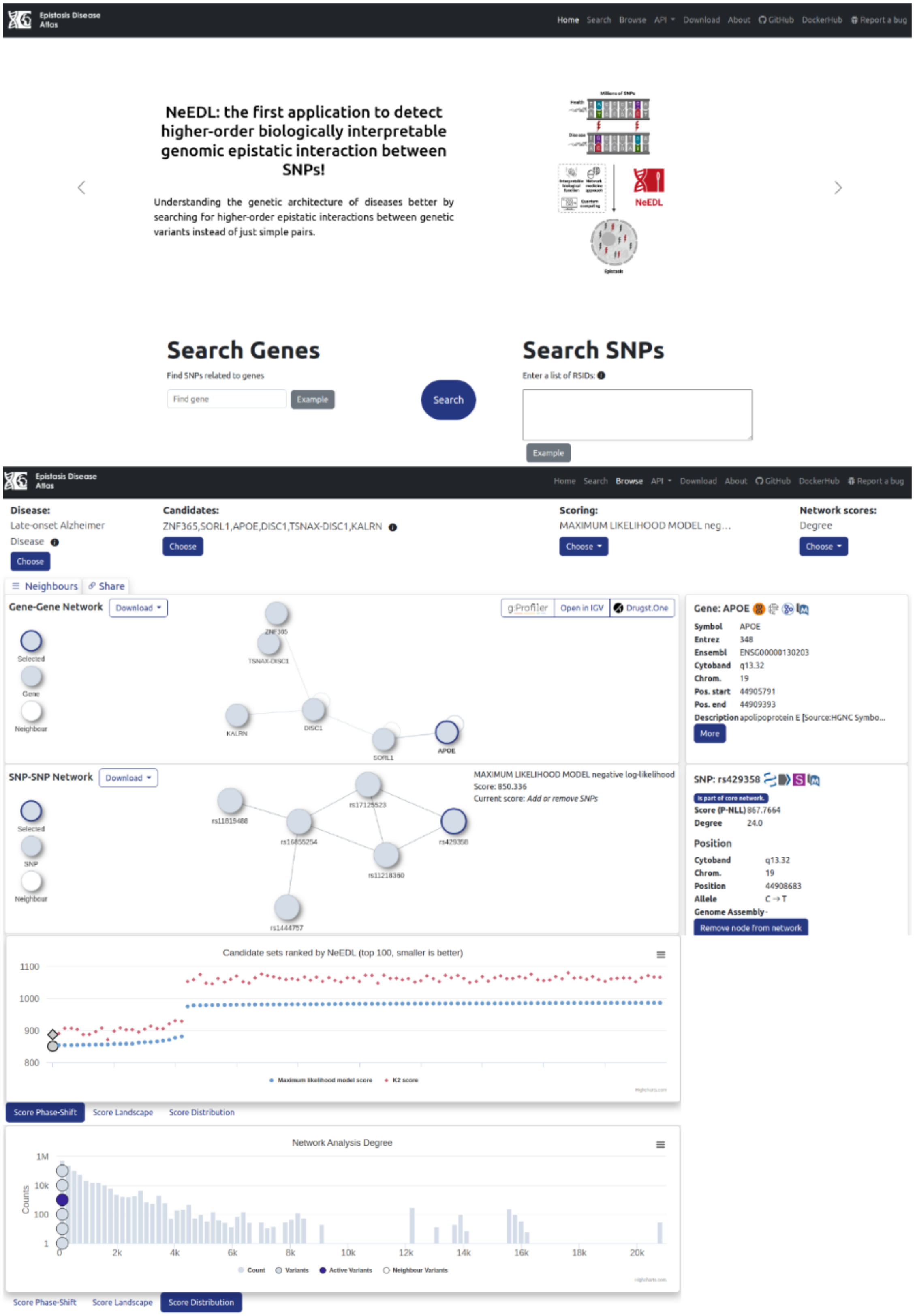


Figure 11: The Epistasis Disease Atlas browse page.

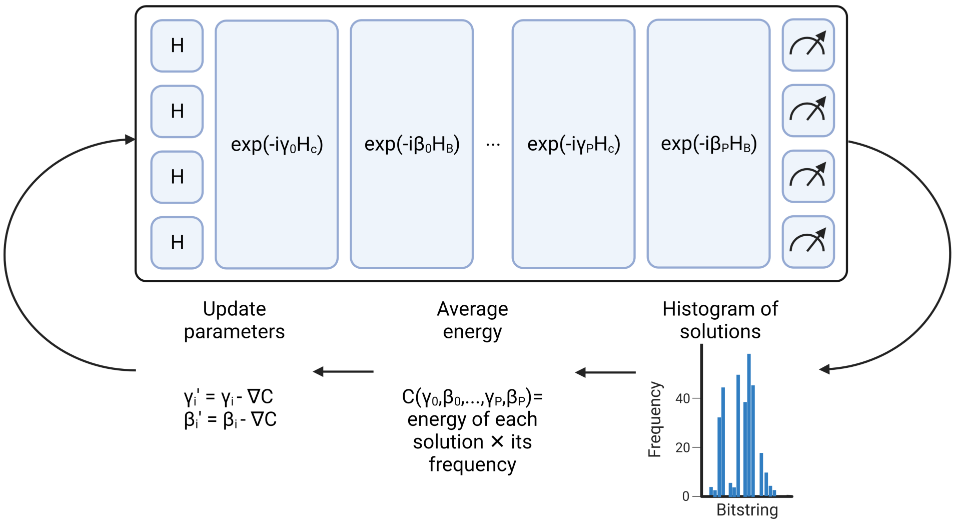


Figure 12: Training of the QAOA circuit. The initial parameters are chosen randomly, and the execution of the circuit samples a histogram of possible solutions. The solutions are then used to define the average cost as the cost of each solution multiplied by their frequency. The cost function is then used to guide the optimization, often by gradient descent.

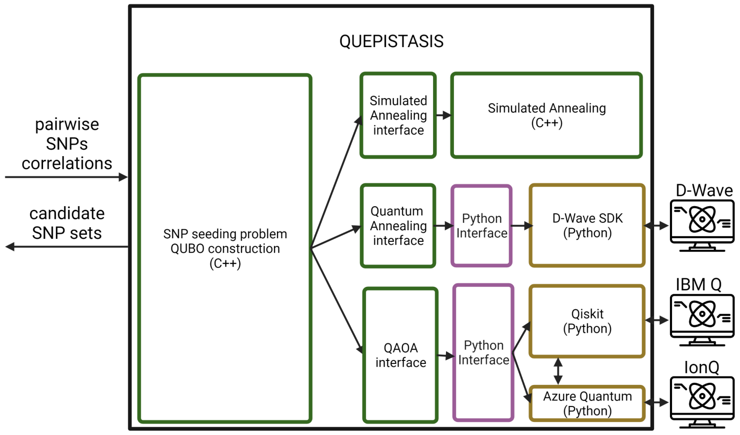


Figure 13: High-level structure of *quepistasis*. The module takes as input a matrix of pairwise relations among SNPs, which is processed by a preliminary software layer to generate a QUBO formulation based on the input data. This QUBO instance can then be solved either by a thermal annealing algorithm on a classical device, or by using one of the available quantum devices such as D-Wave, IBM Quantum devices, or the devices accessible through the Azure Quantum cloud, which includes IonQ devices. The output is one or more SNP sets that will be used as seeds for the local search.

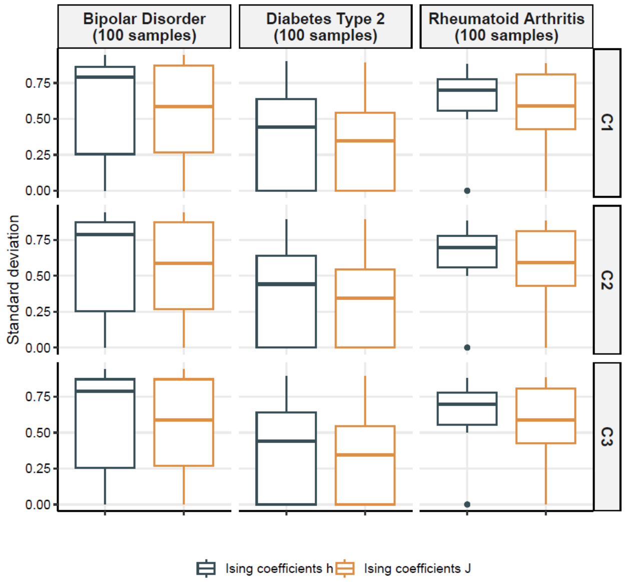


Figure 14: Standard deviation of the Ising coefficients $h$ and $J$ in the various subproblems generated by the community detection tool for the three datasets shown and for some configuration. Legend: C1 is the configuration in Suppl. Table 14 having $\lambda_0 = 10.0, \lambda_1 = 0.0001$, C2 having $\lambda_0 = 10.0, \lambda_1 = 1.0$, and C3 having $\lambda_0 = 2.0, \lambda_1 = 1.0$.

Table 1: Number of cases and SNPs per disease that are discussed in this manuscript before and after the usage of epiJSON.

| disease | Raw data set | | Cleaned and filtered data set | | | |
|---|---|---|---|---|---|---|
| | SNPs | samples | SNPs | cases | controls | sum of samples |
| LOAD | 759916 | 1599 | 6041 | 997 | 573 | 1570 |
| BD | 500568 | 3502 | 142441 | 1993 | 1498 | 3491 |
| CAD | 500568 | 3492 | 142347 | 1979 | 1498 | 3477 |
| T1D | 500568 | 3504 | 142432 | 1997 | 1498 | 3495 |
| T2D | 500568 | 3503 | 142546 | 1973 | 1498 | 3471 |
| HT | 500568 | 3505 | 142272 | 1996 | 1498 | 3494 |
| IBD | 500568 | 3509 | 142879 | 1993 | 1498 | 3491 |
| RA | 500568 | 3503 | 142466 | 1973 | 1498 | 3471 |

Table 2: Effects of the filters of epiJSON (pre-filtering and pre-cleaning steps) on numbers of SNPs and samples in Late-Onset Alzheimer's Disease (LOAD).

| filter step | #SNPs | #samples | #cases | #controls |
|---|---|---|---|---|
| raw data | 759916 | 1599 | 1014 | 585 |
| merge case and control | control were already included | | | |
| RS-ID and duplicate filter | 756940 | 1599 | 1014 | 585 |
| missing phenotype | 756940 | 1599 | 1014 | 585 |
| missingness >20% | 756940 | 1599 | 1014 | 585 |
| sex discrepancy | 756940 | 1570 | 997 | 573 |
| MAF <5% | 756940 | 1570 | 997 | 573 |
| HWE | 756940 | 1570 | 997 | 573 |
| heterozygosity | 756940 | 1570 | 997 | 573 |
| missingness >0% | 17044 | 1570 | 997 | 573 |
| dbSNP database | 9550 | 1570 | 997 | 573 |
| SNP-SNP connectivity | 6041 | 1570 | 997 | 573 |

Table 3: Effects of the filters of epiJSON (pre-filtering and pre-cleaning steps) on numbers of SNPs and samples in Bipolar Disorder (BD).

| filter step | #SNPs | #samples | #cases | #controls |
|---|---|---|---|---|
| raw data | 500568 | 3502 | 1998 | 1504 |
| merge case and control | 500566 | 3502 | 1998 | 1504 |
| RS-ID and duplicate filter | 500566 | 3502 | 1998 | 1504 |
| missing phenotype | 500566 | 3502 | 1998 | 1504 |
| missingness >20% | 500566 | 3502 | 1998 | 1504 |
| sex discrepancy | 500566 | 3491 | 1993 | 1498 |
| MAF <5% | 384955 | 3491 | 1993 | 1498 |
| HWE | 379007 | 3491 | 1993 | 1498 |
| heterozygosity | 379007 | 3491 | 1993 | 1498 |
| missingness >0% | 379007 | 3491 | 1993 | 1498 |
| dbSNP database | 207273 | 3491 | 1993 | 1498 |
| SNP-SNP connectivity | 142441 | 3491 | 1993 | 1498 |

Table 4: Effects of the filters of epiJSON (pre-filtering and pre-cleaning steps) on numbers of SNPs and samples in Coronary Artery Disease (CAD).

| filter step | #SNPs | #samples | #cases | #controls |
|---|---|---|---|---|
| raw data | 500568 | 3492 | 1988 | 1504 |
| merge case and control | 500566 | 3492 | 1988 | 1504 |
| RS-ID and duplicate filter | 500566 | 3492 | 1988 | 1504 |
| missing phenotype | 500566 | 3492 | 1988 | 1504 |
| missingness >20% | 500566 | 3492 | 1988 | 1504 |
| sex discrepancy | 500566 | 3477 | 1979 | 1498 |
| MAF <5% | 384643 | 3477 | 1979 | 1498 |
| HWE | 378631 | 3477 | 1979 | 1498 |
| heterozygosity | 378631 | 3477 | 1979 | 1498 |
| missingness >0% | 378631 | 3477 | 1979 | 1498 |
| dbSNP database | 207105 | 3477 | 1979 | 1498 |
| SNP-SNP connectivity | 142347 | 3477 | 1979 | 1498 |

Table 5: Effects of the filters of epiJSON (pre-filtering and pre-cleaning steps) on numbers of SNPs and samples in Diabetes Type 1 (T1D).

| (d) Diabetes Type 1 (T1D) | | | | |
|---|---|---|---|---|
| filter step | #SNPs | #samples | #cases | #controls |
| raw data | 500568 | 3504 | 2000 | 1504 |
| merge case and control | 500566 | 3504 | 2000 | 1504 |
| RS-ID and duplicate filter | 500566 | 3504 | 2000 | 1504 |
| missing phenotype | 500566 | 3504 | 2000 | 1504 |
| missingness >20% | 500566 | 3504 | 2000 | 1504 |
| sex discrepancy | 500566 | 3495 | 1997 | 1498 |
| MAF <5% | 384838 | 3495 | 1997 | 1498 |
| HWE | 378856 | 3495 | 1997 | 1498 |
| heterozygosity | 378856 | 3495 | 1997 | 1498 |
| missingness >0% | 378856 | 3495 | 1997 | 1498 |
| dbSNP database | 207255 | 3495 | 1997 | 1498 |
| SNP-SNP connectivity | 142432 | 3495 | 1997 | 1498 |

Table 6: Effects of the filters of epiJSON (pre-filtering and pre-cleaning steps) on numbers of SNPs and samples in Diabetes Type 2 (T2D).

| filter step | #SNPs | #samples | #cases | #controls |
|---|---|---|---|---|
| raw data | 500568 | 3503 | 1999 | 1504 |
| merge case and control | 500566 | 3503 | 1999 | 1504 |
| RS-ID and duplicate filter | 500566 | 3503 | 1999 | 1504 |
| missing phenotype | 500566 | 3503 | 1999 | 1504 |
| missingness >20% | 500566 | 3503 | 1999 | 1504 |
| sex discrepancy | 500566 | 3471 | 1973 | 1498 |
| MAF <5% | 384967 | 3471 | 1973 | 1498 |
| HWE | 379211 | 3471 | 1973 | 1498 |
| heterozygosity | 379211 | 3471 | 1973 | 1498 |
| missingness >0% | 379211 | 3471 | 1973 | 1498 |
| dbSNP database | 207379 | 3471 | 1973 | 1498 |
| SNP-SNP connectivity | 142546 | 3471 | 1973 | 1498 |

Table 7: Effects of the filters of epiJSON (pre-filtering and pre-cleaning steps) on numbers of SNPs and samples in Hypertension (HT).

| filter step | #SNPs | #samples | #cases | #controls |
|---|---|---|---|---|
| raw data | 500568 | 3505 | 2001 | 1504 |
| merge case and control | 500566 | 3505 | 2001 | 1504 |
| RS-ID and duplicate filter | 500566 | 3505 | 2001 | 1504 |
| missing phenotype | 500566 | 3505 | 2001 | 1504 |
| missingness >20% | 500566 | 3505 | 2001 | 1504 |
| sex discrepancy | 500566 | 3494 | 1996 | 1498 |
| MAF <5% | 384400 | 3494 | 1996 | 1498 |
| HWE | 378553 | 3494 | 1996 | 1498 |
| heterozygosity | 378553 | 3494 | 1996 | 1498 |
| missingness >0% | 378553 | 3494 | 1996 | 1498 |
| dbSNP database | 207051 | 3494 | 1996 | 1498 |
| SNP-SNP connectivity | 142272 | 3494 | 1996 | 1498 |

Table 8: Effects of the filters of epiJSON (pre-filtering and pre-cleaning steps) on numbers of SNPs and samples in Inflammatory Bowel Disease (IBD).

| filter step | #SNPs | #samples | #cases | #controls |
|---|---|---|---|---|
| raw data | 500568 | 3509 | 2005 | 1504 |
| merge case and control | 500566 | 3509 | 2005 | 1504 |
| RS-ID and duplicate filter | 500566 | 3509 | 2005 | 1504 |
| missing phenotype | 500566 | 3509 | 2005 | 1504 |
| missingness >20% | 500566 | 3509 | 2005 | 1504 |
| sex discrepancy | 500566 | 3491 | 1993 | 1498 |
| MAF <5% | 386025 | 3491 | 1993 | 1498 |
| HWE | 380215 | 3491 | 1993 | 1498 |
| heterozygosity | 380215 | 3491 | 1993 | 1498 |
| missingness >0% | 380215 | 3491 | 1993 | 1498 |
| dbSNP database | 207890 | 3491 | 1993 | 1498 |
| SNP-SNP connectivity | 142879 | 3491 | 1993 | 1498 |

Table 9: Effects of the filters of epiJSON (pre-filtering and pre-cleaning steps) on numbers of SNPs and samples in Rheumatoid Arthritis (RA).

| filter step | #SNPs | #samples | #cases | #controls |
|---|---|---|---|---|
| raw data | 500568 | 3503 | 1999 | 1504 |
| merge case and control | 500566 | 3503 | 1999 | 1504 |
| RS-ID and duplicate filter | 500566 | 3503 | 1999 | 1504 |
| missing phenotype | 500566 | 3503 | 1999 | 1504 |
| missingness >20% | 500566 | 3503 | 1999 | 1504 |
| sex discrepancy | 500566 | 3471 | 1973 | 1498 |
| MAF <5% | 384923 | 3471 | 1973 | 1498 |
| HWE | 379039 | 3471 | 1973 | 1498 |
| heterozygosity | 379039 | 3471 | 1973 | 1498 |
| missingness >0% | 379039 | 3471 | 1973 | 1498 |
| dbSNP database | 207314 | 3471 | 1973 | 1498 |
| SNP-SNP connectivity | 142466 | 3471 | 1973 | 1498 |

Table 10: Feature comparison between NeEDL and existing epistasis detection tools which we initially aimed to include in our benchmark. We selected recently published methods and methods that were incorporated into recently published methods (LinDen into Potpourri). Due to the unresolvable problems summarized in Suppl. Table 11, the benchmark was ultimately run with MACOED and LinDen as the only competitors.

| | **NeEDL** | **PoCos** | **MACOED** | **LinDen** | **Potpourri** | **biologicalEpistasis** |
|---|---|---|---|---|---|---|
| **random subroutines** | start seeds selection | none | ant colony optimization | none | none | none |
| **statistical models** | $\chi^2$-test, multiple quadratic regression scores, Bayesian K2-score, maximum likelihood model | quadratic regression | $\chi^2$-test, Akaike information criterion quadratic regression, Bayesian K2-score | $\chi^2$-test | $\chi^2$-test | Regr. for SNP interactions, truncated product method to derive pair $P$-value, simulated distribution with permutations under the null hypothesis |
| **candidate SNP set size** | variable (default: 2-10) | hundreds to thousands | 2 | 2 | 2 | 2 |
| **implementation** | C++ using boost and iGraph | MatLab | C++ | C++ | C++/ MatLab | nextflow pipeline using R and bash |

Table 11: Software problems encountered during benchmarking. Tools, where some problems could not be resolved after months of debugging, were excluded from the benchmark.

| Tool | Problem | Resolved |
|---|---|---|
| PoCos | reported SNP sets are too big to be statistically evaluated (size > 2000 SNPs) | **not resolvable** |
| PoCos | the program does not report the found candidate SNP sets out of the box | resolved |
| PoCos | implemented in the proprietary language MatLab which complicates deploying it on a cluster | resolved |
| MACOED | configuration is not correctly parsed if entries are too long (buffer overflow potentially overwrites internal data structure) | resolved |
| MACOED | exhaustive memory consumption (1.3 TB) | resolved |
| MACOED | overwrites its own results | resolved |
| MACOED | no build dependencies | resolved |
| MACOED | no build pipeline available (e.g., CMake) | resolved |
| MACOED | computationally not parallelizable | **not resolved, but not necessary for usage** |
| Potpourri | crashes on input data without useful error messages (segmentation fault) despite correct formatting after their instructions and same format as example input data | **not resolvable** |
| Potpourri | user needs to write his/her own Matlab script alongside a demo script to run on custom data | resolved |
| Potpourri | is implemented in the proprietary language MatLab which complicates deploying it on a cluster | resolved |
| biologicalEpistasis | cannot be run on larger HPC clusters because the pipeline spawns slurm jobs exhaustively | **not resolvable** |
| biologicalEpistasis | did not terminate a single dataset after 3 months of runtime | **not resolvable** |

Table 12: Computational resources used for our benchmark study. Note that MACOED was run 100 times for each benchmark dataset to account for the fact that it contains randomized subroutines.

| Tool | Main memory (estimates) | total runtime per benchmark dataset (estimates) |
|---|---|---|
| NeEDL | 50 GB | 2 hours |
| LinDen | 500 MB | 0.5 hours |
| MACOED | 1.3 TB | 48 hours |

Table 13: Data from UK Biobank (www.ukbiobank.ac.uk, project IDs 32683 and 54273) used for running replication in Late-Onset Alzheimer's Disease (LOAD). Individuals were selected based on ICD-10 code inclusion, controls were sampled excluding individuals with any of the ICD-10 code exclusion.

| data set | #case/control | ICD-10 code inclusion | ICD-10 code exclusion |
|---|---|---|---|
| LOAD British ancestry | 758/8055 | G30 | G, E, F |
| LOAD mixed ancestry | 845/8954 | G30 | G, E, F |

Table 14: Table of parameters used for conducting experiments on NeEDL via the quantum computing seeding procedure. Legend: values $c_{\min}$ and $c_{\max}$ denote the minimum and maximum size of the subproblem, which the community detection tool utilizes to divide large instances into smaller ones. All other parameters are related to the QUBO formulation. Specifically, $K$ represents the desired size of the SNPs set; $N$ indicates the number of SNP sets returned; $\nu$ denotes the weight of the linear combination of statistical and biological information in the pairwise correlation metric between SNPs; $\lambda_0, \lambda_1, \lambda_2$ denote the weight of the different terms in the QUBO formulation.

| Device | $c_{\min}$ | $c_{\max}$ | $K$ | $N$ | $\nu$ | $\lambda_0$ | $\lambda_1$ | $\lambda_2$ |
|---|---|---|---|---|---|---|---|---|
| Simulated Annealing | 2 | 10 | 5 | 1 | 0.5 | 5.0 | 1.0000 | 1.0000 |
| Simulated Annealing | 2 | 10 | 5 | 1 | 0.5 | 10.0 | 2.0000 | 1.0000 |
| Simulated Annealing | 2 | 10 | 5 | 1 | 0.5 | 10.0 | 1.0000 | 2.0000 |
| Simulated Annealing | 2 | 10 | 5 | 1 | 0.5 | 2.0 | 1.0000 | 1.0000 |
| Simulated Annealing | 2 | 100 | 5 | 1 | 0.5 | 5.0 | 1.0000 | 1.0000 |
| Simulated Annealing | 2 | 100 | 5 | 1 | 0.5 | 10.0 | 2.0000 | 1.0000 |
| Simulated Annealing | 2 | 100 | 5 | 1 | 0.5 | 10.0 | 1.0000 | 2.0000 |
| Simulated Annealing | 2 | 100 | 5 | 1 | 0.5 | 2.0 | 1.0000 | 1.0000 |
| Simulated Annealing | 2 | 100 | 5 | 1 | 0.5 | 10.0 | 0.1000 | 0.1000 |
| Simulated Annealing | 2 | 100 | 5 | 1 | 0.5 | 10.0 | 0.0500 | 0.0500 |
| Simulated Annealing | 2 | 100 | 5 | 1 | 0.5 | 10.0 | 0.0100 | 0.0100 |
| Simulated Annealing | 2 | 100 | 5 | 1 | 0.5 | 10.0 | 0.0010 | 0.0010 |
| Simulated Annealing | 2 | 100 | 5 | 1 | 0.5 | 10.0 | 0.0050 | 0.0050 |
| Simulated Annealing | 2 | 100 | 5 | 1 | 0.5 | 10.0 | 0.0010 | 0.0010 |
| Simulated Annealing | 2 | 100 | 5 | 1 | 0.5 | 10.0 | 0.0005 | 0.0005 |
| Simulated Annealing | 2 | 100 | 5 | 1 | 0.5 | 10.0 | 0.0001 | 0.0001 |
| D-Wave Advantage 6.1 | 2 | 100 | 5 | 1 | 0.5 | 5.0 | 1.0000 | 1.0000 |
| D-Wave Advantage 6.1 | 2 | 100 | 5 | 1 | 0.5 | 10.0 | 2.0000 | 1.0000 |
| D-Wave Advantage 6.1 | 2 | 100 | 5 | 1 | 0.5 | 10.0 | 1.0000 | 2.0000 |
| D-Wave Advantage 6.1 | 2 | 100 | 5 | 1 | 0.5 | 2.0 | 1.0000 | 1.0000 |
| D-Wave Advantage 6.1 | 2 | 100 | 5 | 1 | 0.5 | 10.0 | 0.0000 | 0.0000 |
| D-Wave Advantage 6.1 | 2 | 100 | 5 | 1 | 0.5 | 10.0 | 0.0100 | 0.0100 |
| D-Wave Advantage 6.1 | 2 | 100 | 5 | 1 | 0.5 | 10.0 | 0.0050 | 0.0050 |
| D-Wave Advantage 6.1 | 2 | 100 | 5 | 1 | 0.5 | 10.0 | 0.0010 | 0.0010 |
| QAOA (simulated) | 2 | 5 | 5 | 1 | 0.5 | 5.0 | 1.0000 | 1.0000 |
| QAOA (simulated) | 2 | 5 | 5 | 1 | 0.5 | 10.0 | 2.0000 | 1.0000 |
| QAOA (simulated) | 2 | 5 | 5 | 1 | 0.5 | 10.0 | 1.0000 | 2.0000 |
| QAOA (simulated) | 2 | 5 | 5 | 1 | 0.5 | 2.0 | 1.0000 | 1.0000 |
| QAOA (simulated) | 2 | 5 | 5 | 1 | 0.5 | 10.0 | 0.0100 | 0.0100 |
| QAOA (simulated) | 2 | 5 | 5 | 1 | 0.5 | 10.0 | 0.0050 | 0.0050 |
| QAOA (simulated) | 2 | 5 | 5 | 1 | 0.5 | 10.0 | 0.0010 | 0.0010 |
| QAOA (simulated) | 2 | 5 | 5 | 1 | 0.5 | 10.0 | 0.0001 | 0.0001 |
| QAOA (IBM) | 2 | 5 | 5 | 1 | 0.5 | 10.0 | 0.0005 | 0.0005 |

### Supplementary Materials

#### Supplementary Materials 1: Biological plausibility studies for Diabetes Type 1 and Inflammatory Bowel Disease

For T1D, a gene associated with 19 high-prevalence SNP combinations was SRPK2. SPRK2 is involved in the regulation of mTORC-1-driven metabolic disorders, which includes T1D[20,21]. NeEDL identified 23 additional genes exhibiting potential epistatic SNP combinations with SPRK2 (Suppl. File 2). GSEA determined that 9 of these genes (NCK1, ANK3, DAW1, BTRC, CHD1L, NRG1, ERBB3, HLA-DPB1, RHBDD1) in addition to SRPK2 were significantly enriched in protein binding activity (GO:0030674; k/K= 0.0144; $P$-value= $9.99e^{-07}$; FDR $q$-value= $7.24e^{-03}$) or boosting gene expression (GO:0010628; k/K= 0.006; $P$-value= $1.83e^{-06}$; FDR $q$-value=$7.24e^{-03}$). One of these genes is NCK1, which is known to impact insulin biogenesis in pancreatic beta cells as well as their survival[22,23]. Another one is NRG1, which is reported to regulate glucose tolerance[24]. We also identified ERBB3, which has been well recognized for its association with T1D risk[25–28].

For IBD, the most prevalent gene present in the top 42 candidate SNP sets is SOX5. A transcription factor analysis of the IBD-associated mucosal transcriptome showed SOX5 binding motifs were the most prevalent inflammation-associated sites for Crohn's Disease (CD) and Ulcerative Colitis (UC) patients[29]. NeEDL identified 42 additional genes exhibiting potential epistatic SNP combinations with SOX5 (Suppl. File 2). GSEA determined that 10 of these genes (TAF6, SMAD1, TAF3, KAT5, ESR1, MED27, SSBP3, LMO2, PASD1, DACH1) were significantly enriched in a gene set that is involved in building a protein complex that controls DNA transcription (GO:0005667; k/K=0.02; $P$-value= $3.13e^{-11}$; FDR $q$-value= $3.31e^{-07}$) with all but TAF3 significantly enriched along with SOX5, SOX6, and CREB5 in a gene set in the molecular function that regulates gene transcription for proper expression in cells and organisms (GO:0140110; k/K=0.0062; $P$-value= $1.34e^{-07}$; FDR $q$-value= $2.36e^{-04}$).

#### Supplementary Materials 2: Epistasis Disease Atlas

To make NeEDL available to the community and allow user-driven research of epistasis, we present the Epistasis Disease Atlas (`https://epistasis-disease-atlas.com/home`). It is a whole platform enabling users to browse precomputed findings within 8 human heritable diseases (see Section Datasets). We further offer real-time on-demand recomputation of epistasis scores for user-defined variant sets. The Epistasis Disease Atlas is an integrative resource providing filters, visualizations, and additional, relevant information about variants, genes, and pathways linking to widely used resources.

##### Database

For storage, a relational database (MySQL) and a graph database (Neo4j) are used in combination to get the best out of both systems. Any large n:m mapping between different entities, like "which genes are affected by a variant" or "which genes products interact with each other", can be represented in the Neo4j instance and efficiently queried. The SQL database stores any other annotated information like common names for diseases, other protein IDs, or pathway descriptions. A complete UML representation depicting the database structures can be found in Suppl. Figure 9 and Suppl. Figure 10, respectively. The sources of the stored information can be separated into two categories. Generated or computed information is everything that is connected to executing NeEDL, this includes the evaluated datasets, runs, candidate sets, and scores. The second type is integrated data from 5 other sources with the goal of providing additional layers of information to the Epistasis Disease Atlas user. We further integrate the Human Protein Atlas (HPA)[30] to provide gene expression information of relevant tissues, WikiPathways[31] to show associated pathways, and Gene Ontology (GO)[32] to show GO classification for selected genes. The MV-Physical dataset of BioGrid[33] provides a protein interaction network, translated to a gene interaction network for network visualization. Variant rsID and location information from dbSNP of UCSC[34](release 155) is imported using BigBed tools[35] and extended by allele and impact information using ClinVar data[36] (release 2023-03-05) and cytoband annotation from UCSC[34] (cytoband.txt.gz). For the majority of the data, the import process is automatized; solely the list of tissues affected by the disease investigated in a dataset was manually curated by conducting a literature search.

##### Application programming interface (API)

The API server of the Epistasis Disease Atlas platform manages access to the databases, handles the asynchronous on-demand execution of the NeEDL algorithm, and makes results on the website reloadable. First-level applications like the website but also the R and Python packages utilize the API to provide functionalities, but documentation for the Epistasis Disease Atlas API is provided to be used by anyone in need of more control and functions (`https://epistasis-disease-atlas.com/api/schema/redoc/`, `https://epistasis-disease-atlas.com/api/schema/swagger/`).

All server applications are deployed using docker; the API is written in Django (v4.1.9) for Python 3.9, providing a RESTful API using djangorestframework (v3.13.1). An OpenAPI 3-based library called drf-spectacular (v0.23.1) is used to render swagger and redoc documentation. Databases are MySQL (v5.7), and Neo4j (v4.4-enterprise), and queries are executed using mysqlclient (v2.1.0) and neomodel (v3.0.8) modules. Redis is used as the broker for the RabbitMQ queue handling tasks for Celery (v5.2.2) workers allowing execution of tasks in the background.

##### File download server, R and Python packages

We offer the full results calculated by NeEDL of 8 human heritable diseases (see Section Datasets) as a zip download via a static file server (https://github.com/halverneus/static-file-server) under https://files.epistasis-disease-atlas.com/. For convenience purposes for the user, we offer an R and Python package to directly load the results of NeEDL into the programming environment. The R and Python package can directly interact with the API of the Epistasis Disease Atlas and thus offers most data that is used for visualization in the web application.

##### Feature-rich web application

The main entry point for users, interested in data from the Epistasis Disease Atlas, is the web frontend. The website focuses on usability for researchers exploring epistasis research questions. Any precomputed results from available datasets are published there (see Section Datasets). There are three different ways to start the exploration of epistasis disease data: A query can be started by searching either for key genes or variants that should be investigated because of their hypothesized epistatic effects, leading to a list of candidate sets and thus runs these entities participated in, from which on entry can be selected. A third alternative is to start browsing the precomputed data directly. The browser page (https://epistasis-disease-atlas.com/browse) requires the selection of the dataset before listing all pre-computed sets of variants with the respective result scores. In all cases, upon selection, the result page for the given run is loaded, and results can be further explored, or on-demand scores can be requested.

In the result explorer page (Suppl. Figure 11), selected genes are integrated with the interactome (inferred from protein-protein interactions). Upon selecting a run, the respective variant (SNP-SNP) interaction network is constructed, as well as the energy landscape determined in the said run. Results are not just immutable, but functions are provided to add additional variants from the interaction neighborhood to investigate new epistasis hypotheses and expand on precalculated results. Candidate SNPs can be filtered by network properties in the variant interaction network and other scores to limit and only investigate relevant options. Recalculation of updated variant sets can be done live without great delay. Researched variant sets are internally associated with a unique identifier encoded in the explorer page URL. This allows results to be shared with fellow researchers or saved to a document to regenerate the exact same result page.

The platform cares about integrating into a researcher's common way of exploration by giving additional information about genes, variants, associated pathways, and go classification, as well as link-outs to popular follow-up analysis tools like g:Profiler or Drugst.One[37]. For custom follow-up analysis, downloads (results in CSV format and networks as an SQLite file) for different parts of the analyzed variant set are provided.

The platform is designed to be easily updateable once more datasets become available and are analyzed by the NeEDL Algorithm.

The website was built with npm (v9.1.2), is written in AngularJS (v13.1.0) with typescript (v4.5.2) for node (v12.20.55), and styled with bootstrap (v5.1.3). For data visualization, vis.js (vis-data: v7.1.4, vis-network v9.1.2) is used for networks, datatables.net (v1.13.1) for tables, and highcharts (v10.1.0) for diagrams.

#### The Integrated Genome Browser plugin

To visualize SNPs in potentially important regions on the genome, we integrated an enhanced genome browser plug-in which includes the dependencies: npm 0.2.0, angular v13.1.0, typescript v4.5.2, Design: angular/material v13.3.9, Plots: @swimlane/ngx-charts v20.4.1, igv v2.15.8[38–41], drugs.one v1.1.17[37].

This plugin includes the integration of the Human Protein Atlas[42], GeneOntology[43], GeneCards[44], WikiPathways[31], g:Profiler[45], Drugst.one[37], the Integrated Genome Browser[38–41], ChIP-Atlas[46,47], ReMap2022[48], ENA[49], ENCODE[50], GEO[51], JASPAR2022[52], EPDnew[53,54], and Enhancer Atlas 2.0[55] for further seamless analysis.

##### Automatically generated R Shiny App that incorporates most functionalities of the Epistasis Disease Atlas for datasets that are processed on local machines or clusters

The Epistasis Disease Atlas can only visualize results and offer its functionalities for datasets that were preprocessed and stored in its database. However, GWAS data is mostly protected by strict licenses and cannot be shared, hence, we offer an R Shiny app that is automatically built after NeEDL successfully finished its analyses for data that is locally processed at the users' machine or cluster. The R Shiny app offers most functions of the Epistasis Disease Atlas for the convenience of the user. The R Shiny App is built on the following R Shiny packages (shiny v1.7.4, shinyWidgets v0.7.6, shinydashboard v0.7.2, shinyjs v2.1.0, and shinycssloaders v1.0.0).

#### Supplementary Materials 3: Functioning of quantum gate-based hardware

NeEDL is capable of interfacing with various quantum circuit-based hardware through its interface with both the Microsoft Azure Quantum Cloud and IBM Quantum platform. The Microsoft Azure Quantum Cloud provides access to ion-trap-based quantum processing units (QPUs) from IonQ, as well as other types of quantum devices added recently. The IBM Quantum platform offers access to a variety of superconducting-based QPUs. The hardware devices are briefly described to provide an overview of their respective features.

##### Ion-trap-based devices: IonQ Harmony

The IonQ Harmony quantum processor unit has a complete graph topology with 11 fully connected qubits, providing a significant advantage over superconducting-based QPUs that have limited sparse connectivity and require substantial overhead to operate on distant qubits. For detailed information on the technical specifications and performance benchmarks of IonQ devices, we refer to[56]. Additionally, information about the specific native gate set and rotation precision of IonQ's Harmony device can be found in[57] and information about their system fidelity can be found on Microsoft Azure Documentation which provided access to these machines[58].

The IonQ Harmony Quantum Processing Unit operates using a native gate set composed of single-qubit gates $GPI(\phi)$, $GPI2(\phi)$, and $RZ(\phi)$, as well as the two-qubit gate $MS(\phi_1, \phi_2)$. The specific definitions of these gates are shown in[57]. The IonQ Harmony QPU uses a set of native gates that is universal, i.e. can be used to create any unitary and therefore execute any quantum circuit. Careful design of the quantum circuit to follow this native set of gates can reduce the overhead of the compilation process. In particular, the operation $RZ$ is physically more efficiently executed than $GPI$ and $GPI2$ gates and should be favored when possible. For example, a Hamiltonian of $H = \sum_i X_i + \sum_{i,j} Z_i Z_j$ can be reformulated to $H = \sum_{i,j} X_i X_j + \sum_i Z_i$ for optimal execution on the IonQ Harmony QPU. The compiler can perform some of these transformations automatically, but for more complex cases, human intervention may be required.

The gates implemented may not perfectly match their theoretical counterparts. On average, the single-qubit gate fidelity is 99.35% and the two-qubit gate fidelity is 96.02%. Additionally, the rotational gates have a precision of $\pi 10^{-3}$.

The quantum state of the system remains coherent, i.e. it evolves correctly for a time frame that can be estimated using two constants: the $T_1$ relaxation time, which measures the time it takes for a state $|1\rangle$ to decay to state $|0\rangle$, with value $10^7 \mu s$, and the $T_2$ dephasing time, which measures the average time the system loses phase and is $2 \cdot 10^6 \mu s$. These values are much higher compared to superconducting-based quantum hardware, indicating a significant performance advantage in the ion trap-based hardware. However, this advantage is partially worsened by the longer time it takes to perform a quantum operation, which is approximately $10\mu s$ for a single-qubit gate and $200\mu s$ for a two-qubit gate.

**Quantum gate-based hardware: IBMQ Perth and Lagos**

Most of the experiments performed with QAOA are carried out on IBM Quantum devices *Perth* and *Lagos*, which share several attributes such as their physical realization, architecture, and topology (refer to[59] for additional information). These devices have a different technology than the ion-trap-based one, with the former being more conducive to scaling to a large number of qubits, but at the expense of lower fidelity.

The IBM QPUs used in some of our experiments is a superconducting-based quantum processing unit that has 7 qubits arranged in a specific topology. It's important to note that the results of a quantum circuit can vary greatly depending on how the qubits are mapped to the QPU's physical qubits, which is typically done by the software compiler. When qubits are mapped to non-physically adjacent qubits, additional SWAP gates must be added, which incurs a large overhead. A common, although sub-optimal, compiler strategy is to map the qubits with more two-qubit gates applied to them (e.g. CNOTs) to physical qubits with higher connectivity in a sequential, greedy manner. Additionally, it should be noted that some qubits within the architecture may have lower precision compared to others, both in terms of the accuracy of the gates applied to them and the coherence time. The $T_1$ and $T_2$ times have ranges of 24.50 to $132.70\mu s$ and 20.06 to $170.37\mu s$, respectively, whereas the gate time is approximately $0.5\mu s$. The single-qubit and two-qubit fidelity errors have ranges of 1.6 to $6.0 \times 10^{-4}$ and 4.8 to $16.7 \times 10^{-3}$, respectively. The native gate set for both the IBM Perth and Lagos QPUs consists of CNOT, RZ, SX, and X gates, which together form a universal set.

#### Supplementary Materials 4: Functioning of quantum annealers

We describe the physical realization of a quantum annealer, in particular the D-Wave Advantage 6.1. This machine is composed of 5616 superconducting qubits. Each qubit is realized by means of superconducting quantum-interference (SQUID) implementation, that can be controlled via radio frequency pulses[60]. Such pulses are also responsible for and used to manipulate the coupling between the qubits, in such a way that during the annealing time, the system evolves toward the Hamiltonian in which the optimization problem is encoded (see[61] for more details). If the hypothesis of the adiabatic theorem holds, and if the annealing time is sufficiently long, the time-dependent Hamiltonian in Equation (5) will remain in its ground state during the annealing process.

A paramount aspect of quantum annealers is the topology of the QPUs, i.e. how many qubits the machine has and how they are connected to each other. Superconductor-based quantum hardware usually has restrictive topologies, implying that if two non-adjacent qubits must communicate, we will need to pass the information through a chain of qubits connecting them. D-Wave Advantage 6.1 has 40135 couplers to link its 5616 qubit, which largely outperforms its predecessor D-Wave 2000Q in terms of connectivity (with the latter having only 2048 qubits and 6016 couplers). When mapping an instance of QUBO problem onto the quantum hardware, in the best-case scenario, each QUBO variable will be mapped to a single qubit of the QPU. The mapping between QUBO variables to qubits is called *minor embedding* and consists concretely in finding an isomorphism between the graph underlying the QUBO instance and the QPU topology graph. As the best graph isomorphism algorithm's runtime is super-polynomial in the input size[62], we have to rely on heuristics to find a suitable solution. Not only the minor embedding should be efficient in terms of asymptotic complexity but also extremely fast in order to add a negligible overhead to the optimization procedure. The D-Wave software framework, Ocean Software Development Kit, already implements several heuristics to perform minor embedding. A comparison of minor embedding techniques for D-Wave device's topologies is shown in[63]. The degree of the nodes in the topology of D-Wave devices is bounded by some constant, we can expect Ising instances described by dense graphs won't be successfully mapped. However, we can artificially enlarge the connectivity of qubits by grouping several physical qubits into a *chain*, forming a single logical qubit. Then the graph of the Ising problem is mapped to the logical qubits. As the readout process will read all the physical qubits, there is the possibility that the readout finds a solution that has different values for the physical qubits associated with a single logical qubit. This problem, called *chain break*, is resolved using deterministic techniques, such as majority voting, which assigns the most frequent value of the chain to the logical qubit. The resource overhead of implementing chains of physical qubits results in the fact that, if theoretically a quantum annealer with $m$ qubits can solve a QUBO problem on up to $m$ binary variables, in practice, the number is much smaller.

The D-Wave QPU must be programmed using the coefficients $h, J$. Although Ising formulations allow coefficients to range from negative infinity to positive infinity, the quantum annealer requires them to be scaled to a finite range. For the D-Wave Advantage 6.1, the range for $h$ coefficients is $[-4, 4]$ and for $J$ coefficients, it is $[-1, 1]$. It is worth noting that these ranges can be slightly expanded for enforcing specific conditions, such as ensuring adherence to the physical-logical qubit mapping. The encoding precision can impact solution quality especially if there are large coefficients that fall at the extremes of the encoding range, causing smaller coefficients to be scaled to zero near the machine precision. In such a scenario, the D-Wave would effectively be solving a different problem than the one intended. By analyzing the distribution of coefficients $h$ and $J$ we can rule out that our specific instance is affected by the encoding precision problem. Additionally, the encoding precision is nonlinear, with a range of $[0.0005, 0.004]$ for $h$ coefficients and $[0.0005, 0.002]$ for $J$ coefficients, as specified in the technical documentation[64]. This means that certain range coefficients are represented with greater accuracy, suggesting the QUBO formulation may be biased to have most coefficients in such favorable ranges.

The quantum annealer evolves its Hamiltonian according to the equation,

$$H(s) = A(s)H_0 + B(s)H_p \tag{1}$$

where $H_0 = \sum_{i=1}^{n} \sigma_x^{(i)}$ represents the free Hamiltonian, $H_p$ denotes the Hamiltonian of the problem in question, and $s$ is the time variable, normalized to fit within the range $[0, 1]$. The quantum annealer starts by being in the ground state of $H_0$, and as $s$ moves from 0 to 1, $A(s)$ decreases while $B(s)$ increases. At the end of the process, the system is described by $H_p$ and its state may be the ground state of $H_p$. The schedule of the annealer is determined by the equation $A(s)$ and $B(s)$, with $0 \leq A(s) \leq 1$ and $B(s) = 1 - A(s)$. The annealing schedule is defined by the total annealing time in microseconds and the shape of the curve, which is represented by a sequence of points $\{(s_i, p_i) \mid 0 \leq s_i < s_{i+1} \leq 1, 0 \leq p_i \leq p_{i+1} \leq 1 \; \forall i\}$ where $s$ represents the normalized time and $p$ the progress. The schedule can include mid-anneal quenches, which are changes in the slope, and mid-anneal pauses, which have been shown to potentially improve solution quality[65]. Longer annealing times can have both positive and negative effects, as they facilitate the system to remain in the ground state during the whole evolution but also might cause larger decoherence errors.

The operation of a quantum annealer can be affected by multiple sources of noise due to the high number of interacting qubits, leading to a detrimental loss of quantum coherence, known as decoherence. The major sources of noise can be grouped into two categories. The first one, causing dephasing, is related to the interaction of the qubits with the external environment that can be modeled as a $1/f-$like noise. Whereas, the second is due to imperfect tuning of the control fields and of the coupling between first-neighbor qubits that are responsible for effective next-neighbor hopping. In the case of the used quantum annealer, the second type of noise and the induced crosstalks due to imperfect tuning are referred to as *background susceptibility*. Apart from the two major ones, there are also other smaller issues that could affect the execution of a quantum annealer. These include the accuracy of the readout operation, which typically has a fidelity of over 99%, making the error close to negligible. Additionally, there is a low risk of programming errors, with an error rate below 10%.

#### Supplementary Materials 5: Quantum algorithm for SNP seeding

To solve the SNPs seeding problem, which involves finding the optimal set of candidate SNPs for local processing, we construct a SNPs graph, where vertices represent SNPs and edges represent the pairwise correlation (or other metric) between them. We assume that a group of SNPs is a strong candidate if the total pairwise correlation between the SNPs in the group is high. This enables us to model the problem as a *weighted-max-clique* problem, which consists in finding a *clique* with the highest sum of pairwise edge weight. As this is a NP-hard problem, it can be mapped, as explained before, into a QUBO formulation, which is the standard method for implementing combinatorial optimization on quantum hardware.

##### The Max-Clique problem

The Max-Clique problem is the computational problem of finding the maximum clique of an undirected graph and is used to model many real-world problems. A clique on a graph is a complete sub-graph of the given graph, that is, one where all the vertices are connected to all other vertices. To find a maximum clique of a graph, one could non-deterministically first try to determine a set of $k$ distinct vertices and then test whether these vertices form a complete graph. In fact, this problem is NP-Complete, and it is conjectured that no polynomial time deterministic algorithm can be constructed to solve it.

##### Quadratic Unconstrained Binary Optimization (QUBO)

This is a mathematical formulation that can embrace an exceptionally large variety of important combinatorial optimization problems occurring both in industry and in science. The QUBO model has emerged as an underpinning of the quantum computing area known as quantum annealing. It can be shown to be equivalent to the Ising model, which plays a prominent role in physics, as highlighted in the paper by[66], and underlies an important domain of problems arising in physics applications. QUBO models belong to a class of problems known to be NP-hard. Therefore, practically no exact solver designed to find optimal solutions (like the commercial CPLEX and Gurobi solvers) is likely to be successful except for very small problem instances. For our problem, we will use two meta-heuristic methods that are designed to find high-quality but not necessarily optimal solutions, namely *quantum annealing* and *QAOA*.

Mathematically, the QUBO problem requires minimizing a function $f : \{0,1\}^n \to \mathbb{R}$,

$$f(x) = \sum_{i,j=1}^{n} x_i Q_{i,j} x_j \tag{2}$$

where $Q$ is a $n \times n$ real matrix. The QUBO formulation allows $Q$ to be equivalently either symmetrical or upper triangular, as every symmetric matrix is an upper triangular matrix in which the entries have been reflected on the diagonal. The former can be obtained from the latter using the transformation $Q' = Q + Q^T - \text{diag}(Q)$. Note the diagonal terms are linear since $x_i^2 = x_i$. If $Q$ is sparse, we can equivalently describe the problem as a graph $G = (V, E, w : E \to \mathbb{R})$ where $V$ is the set of $n$ binary variables, $E$ the set of edges such that $(u, v) \in E$ if and only if $Q_{u,v} \neq 0$, and $w((u, v)) = Q_{u,v}$. QUBO problem can deal with signed integer and fixed point arithmetic after appropriate encoding on a binary vector; for example, a positive integer can be encoded on $d$ bits on the variable $x \in \{0,1\}^d$ through the term $\sum_{i=0}^{d} 2^i x_i$. Any boolean function acting on $n$ bits can be expressed using an $n$-th degree binary polynomial $f(x) = \sum_{i=1}^{n} a_i x_i + \sum_{i,j=1}^{n} b_i x_i x_j + \sum_{i,j=1}^{n} c_i x_i x_j + \cdots$. Such a formula can be lead back to a quadratic polynomial with the use of auxiliary ('slack') binary variables $\alpha_k = x_i x_j$, although the number of necessary auxiliary variables grows super-polynomially with the degree of the polynomial. QUBO can be used to encode a constrained problem if we encode the constraint as a binary term $C(x)$, such that the formulation can be expressed as $f(x) = \lambda_0 \sum_{i,j=1}^{n} x_i Q_{i,j} x_j + \lambda_1 C(x)$ with $\lambda_0, \lambda_1 > 0$. For $\lambda_0 \ll \lambda_1$, the violation of the constraint results in a large cost $f(x)$, guiding the optimizer toward acceptable solutions. Authors in[67] suggest several strategies to effectively design constraints for QUBO formulations.

##### QUBO formulation for SNPs seeding

We can reduce Max-Clique to QUBO formulation, proving the NP-Harness of the QUBO: given the graph $G = (V = [1, n], E \subseteq V \times V)$ we consider the minimization of the QUBO function over the binary vector $x \in \{0,1\}^n$,

$$f(x) = -\lambda_0 \sum_{i=1}^{n} x_i + \lambda_1 \sum_{(i,j) \notin E} x_i x_j \tag{3}$$

for $\lambda_0 \ll \lambda_1$: the first term reward $x$ vectors with a large number of 1s, corresponding to larger cliques, while the second term penalizes solutions that violate the definition of the clique (vectors containing a vertex not fully connected with the others). The global minima of $f$ is a solution for the max clique problem: the maximum clique is the set $\{v_i \in V \mid x_i = 1\}$.

To formulate epistasis detection as a QUBO problem, we consider a graph $G$ of vertices $V$, edges $E$, and edge weight function $w : V \times V \to \mathbb{R}$ representing the relation between SNPs, i.e. the vertices are the SNPs and between each pair of vertices, there is a positive weight representing the relation between graphs according to some metrics (e.g. correlation).

We guide the optimization through the following assumption: an SNP set of an arbitrary number of elements is a good candidate solution if the sum of the pairwise metric values between SNPs is high.

Thus, the formulation solves and analyzes the graph to find several ($N$) candidate SNP sets, each having an arbitrary size $K$ with the largest sum of edge weights. The formulation is named *the N max-weighted k-cliques.*

$$Q = \sum_{\ell=1}^{N} \left[ \lambda_0 \left( \sum_i x_{i\ell} - K \right) - \lambda_1 \sum_{(i,j)\notin E} w_{i,j} x_{i\ell} x_{j\ell} \right] + \lambda_2 \sum_{\ell=1}^{N} \sum_{m=\ell+1}^{N} \sum_{i=1}^{|V|} x_{i\ell} x_{im} \tag{4}$$

where:

- there are $N \times V$ binary variables $x_{i\ell}$, which is 1 if the $i$-th vertex (SNP) is associated with the $\ell$-th solution set;
- the term $\lambda_0(\sum_i x_{i\ell} - K)$ associate a penalty term for cliques that has a size different than $K$. The penalty is weighed by the hyper-parameter $\lambda_0 > 0$.
- the term $-\lambda_1 \sum_{1\leq i,j\leq |V|} w_{i,j} x_{i\ell} x_{j\ell}$ reward the SNPs set identified with $\{x_{k\ell} : x_{k\ell} = 1, k = 1, ..., |V|\}$ having the largest sum of weights. The penalty is weighed by the hyper-parameter $\lambda_0 > 1$.
- the term $\lambda_2 \sum_{\ell=1}^{n} \sum_{m=\ell+1}^{N} \sum_{i=1}^{|V|} x_{i\ell} x_{im}$ add a penalty for each pair of the $N$ SPS set that are partially or totally overlapping, i.e. are composed of the same SNPs (vertices). The penalty is weighed by the hyper-parameter $\lambda_0 > 2$.

The formulation can be simplified to return $N = 1$ SNPs set and run the optimization technique multiple times with different random initial states. Such a simplification was necessary for any run on the quantum hardware due to the lack of resources.

##### Community detection

Due to quantum computing resource limitations, we could not run NeEDL on datasets containing approx. 140,000 SNPs. Hence, we need to subsample the datasets. We randomly subsample connected subnetworks from the PPI-based SSI network of the data of three diseases, namely, BD, RA, and T2D, to 100, 500, to 1,000 SNPs to showcase the speed-up that quantum computing could give in the future. We use the connected subnetworks within the PPI-derived SSI network and used it as input for the evaluation of the quantum-computing module.

When quantum computing hardware and substantial networks become available, it is not feasible to generate a matrix of all pairwise combinations of SNPs, which is required for quantum computing. Hence, we implemented the community-detection method Leiden[68] to identify more densely connected parts of the network. We calculate the matrix for all pairwise SNP combinations for the subnetwork and use quantum computing on each of the subnetworks. We then choose the most promising regions identified by the quantum computer as starting seeds for the local search. See Suppl. Figure 5b on the right.

##### Supplementary Materials 6: Quantum Algorithms

We briefly review the quantum algorithms that we used to tackle epistasis detection on the currently available quantum computers: Quantum Annealing (QA) and Quantum Approximate Optimization Algorithms (QAOA). Quantum Annealing (QA) is the quantum counterpart to the simulated annealing algorithm[69], a metaheuristic employed to locate an approximate solution to a global optimization problem within a vast search space. Both QA and simulated annealing algorithms operate by identifying the state of minimum energy of spin glass systems studied in statistical mechanics, such as the random field Ising model[70]. The primary distinction between the two methods lies in the adjustable parameter used to introduce the necessary fluctuations for exploring the solution space. In the case of quantum annealing, these fluctuations are generated by a field that permits quantum tunneling, as opposed to simulated annealing, which instead is based on thermal fluctuations[71]. While annealing refers to an adiabatic model of quantum computation, QAOA is a hybrid quantum-classical algorithm that operates on circuit-based quantum computers. Both techniques are based on the total Hamiltonian of the system, $H$, which is a Hermitian operator acting on the Hilbert space $\mathcal{H}$ describing the system of $N$ quantum spins ($\mathcal{H} = \mathbb{C}^{2^N}$). The eigenvalues of this operator correspond to the energy spectrum of the system. After an appropriate mapping of our problem, the strategy is to identify the lowest eigenvalue of the Hamiltonian, which is equivalent to identifying the lowest energy level or ground state of the system. To provide a more rigorous definition, we can describe the general structure of a Hamiltonian. This consists of a free part $H_0$, which accounts for the free energy of all spins in the system, and an interaction term $H_I$, which represents the contribution to the total energy caused by the magnetic interaction among the spins. These two terms are typically non-commuting, i.e., $[H_0, H_1] = H_0H_1 - H_1H_0 \neq 0$, making it extremely difficult to find the exact eigenvalues of $H$ either analytically or numerically, particularly when the number of spins in the system is high, i.e., $N \gg 1$. The total time-dependent Hamiltonian is written as

$$H(t) = (1 - \mu(t))H_0 + \mu(t)H_I, \tag{5}$$

where $\mu(t)$ is a control function that takes values in the interval $[0, 1]$ and allows switching between the two terms. Although each of the two terms could be, in principle, easy to diagonalize, the sum of free terms and interactions could lead to equations that are too complex to be solved exactly. In addition, if the computation time is limited to the interval $[0, \tau]$, a common procedure is to initialize the system in the ground state of $H_0$ with $\mu(0) = 0$, and then adjust the control parameter until reaching $\mu(\tau) = 1$. If the behavior of $\mu(t)$ satisfies the adiabatic theorem[72], both QA and QAOA can be seen as simplified versions of adiabatic quantum computation[73]. As for the QAOA, it is implemented on gate-based quantum hardware by decomposing the unitary evolution $U(t_0, t) = \exp\left\{(-i/\hbar)\int_{t_0}^{t} H(t')dt'\right\}$ from the full Hamiltonian in Equation (5) into the Suzuki-Trotter decomposition[74].

Thermal and Quantum Annealing algorithms[70,75] find their roots in the statistical mechanics' problem of finding the minimum-energy state of a generic Ising model. To explain these techniques, we introduce the random field Ising model, both classical and quantum. For the former we consider classical spin variables $s_i$ that take values $\pm 1$, so that the energy of a system of $N$ interacting spin arranged over a $d-$dimensional lattice is the classical Hamiltonian function:

$$H(s) = -\sum_{\langle i,j \rangle} J_{i,j} s_i s_j - \sum_i h_i s_i. \tag{6}$$

Here the notation $\langle i, j \rangle$ indicates that the sum is over all the pairs of neighboring sites, while the $J_{ij}$ are the couplings strengths between two spins and $h_i$ is a random external field. The quantum version of the Ising Hamiltonian in Equation (6)) is obtained by replacing the binary variable $s_i$ with the corresponding Pauli matrix $\sigma_i^\alpha$, $\alpha = \{x, y, z\}$:

$$H(\sigma) = -\sum_{\langle i,j \rangle} J_{i,j} \sigma_i^z \sigma_j^z - \sum_{i=1}^{N} h_i \sigma^x. \tag{7}$$

The above expression is not anymore a function but a Hermitian operator. It can be linked to the introductory discussion of this section, noting that the free term is $H_0 = \sum_{i=1}^{N} h_i \sigma^x$ and the interaction term coincides with $H_I = -\sum_{\langle i,j \rangle} J_{i,j} \sigma_i^z \sigma_j^z$. This type of Hamiltonian is used to describe the physics of spin glasses and their phases. Finding the minimal energy of such Hamiltonian is a NP-hard problem (specifically NP-complete) and the way it is linked to the solution of many NP problems is reported in[76]. The Ising Hamiltonian formulation is tightly related to some mathematical optimization problems called Quadratic Binary Optimization (QUBO) problems, which belong to the NP-hard complexity class too. A QUBO problem can be mapped to an Ising Hamiltonian by transforming each variable $x_i$ of the former as follows:

$$x_i \leftrightarrow \frac{1 + \sigma_i^z}{2}, \tag{8}$$

This transformation guarantees that the solution of the optimization task will coincide with the search for the ground state of the Ising model considered. We will investigate in the next section the relation between QUBO and the SNP seeding optimization problem. A hardware device dedicated to solving Ising problems is an *Ising machine*. Authors in[77] have reviewed the many Ising machines proposed in the literature, based on different physical phenomena. Quantum annealers such as D-Wave are instances of Ising machines, as well as gate-based quantum hardware when used with the QAOA algorithm.

The second algorithm, QAOA, was introduced in[78] and can be implemented on the currently available noisy intermediate-scale quantum (NISQ) hardware supporting a gate-base model of computation. The algorithm, which is pictured in Suppl. Figure 12, corresponds to a quantum circuit that can be briefly described as follows. The $N-$qubit input state of the quantum circuit is initialized to $|+\rangle^{\otimes N} = (2^{-1/2}|0\rangle + 2^{-1/2}|1\rangle)^{\otimes N}$, where $|+\rangle$ is the eigenvector relative to the eigenvalue 1 of the Pauli matrix $\sigma_x$. Subsequently, a parametrized wavefunction $|\psi(\vec{\gamma}, \vec{\beta})\rangle$ is constructed by designing a suitable ansatz that takes into account the nature of the optimization problem one has to solve. The ansatz is constructed by acting on the initial state $|+\rangle^{\otimes N}$ with $p$ repetitions of a sequence of two parametric unitary operation $U_C(\gamma_k), U_B(\beta_k)$. Therefore, the whole ansatz and the wavefunction will both depend on $2p$ parameters $\gamma_1, \beta_1, ..., \gamma_{2p}, \beta_{2p} \in [-\pi, \pi]$. The link with the Hamiltonian operator defined in Equation (7) is given by setting $\mu(t) = \frac{1}{2}$, then the first unitary operator reads $U_C(\gamma) = \exp\{-i\gamma H_I\}$ and $H_I$ is the term in which the problem is encoded. Lastly, the second unitary operator is defined as $U_B(\beta) = \exp\{-i\beta H_0\}$. We observe that two Hamiltonian operators $H_C, H_B$ (and consequently the associated unitary operators) do not have to commute to obtain meaningful results. Now, we can write the parameterized wavefunction, resulting from the circuit as

$$\left|\psi(\vec{\gamma}, \vec{\beta})\right\rangle = U_B(\beta_p)U_C(\gamma_p)\cdots U_B(\beta_1)U_C(\gamma_1)|+\rangle^{\otimes n}, \tag{9}$$

on which we compute the mean value of the energy of the system defined as

$$E(\vec{\gamma}, \vec{\beta}) = \left\langle \psi(\vec{\gamma}, \vec{\beta}) \middle| H_C \middle| \psi(\vec{\gamma}, \vec{\beta}) \right\rangle. \tag{10}$$

This expression is now a classical function that depends on $2p$ parameters and has to be minimized by means of classical optimization to find the optimal variational parameters $(\vec{\gamma}^*, \vec{\beta}^*)$ such that:

$$(\vec{\gamma}^*, \vec{\beta}^*) = \arg\min_{\vec{\gamma}, \vec{\beta}} E(\vec{\gamma}, \vec{\beta}). \tag{11}$$

#### Supplementary Materials 7: *quepistasis* module

The purpose of the *quepistasis* module is to connect the NeEDL software with the quantum hardware. It requires the input of a pairwise correlation matrix between SNPs, where the correlation can be measured using statistical correlation or any other metric. The output is one or more sets of SNPs, which are then used in the local search procedure. The structure of *quepistasis* is schematized in Suppl. Figure 13.

The *quepistasis* module creates the QUBO formulation of the SNP seeding problem, which can be solved using the classical solver (Simulated Annealing). Since Python 3 is the standard programming language for quantum computing and supports most quantum SDKs, we have written an interface between NeEDL and this software. Thus, the Python interface submodule takes the QUBO matrix and the target platform as inputs, along with some configuration parameters, and returns the binary vector solution found by the quantum device. This solution is then interpreted as an SNP set.

It is important to notice that the *quepistasis* is supported by the community detection tool: as the D-Wave quantum annealer and IBM/IonQ devices can deal with problems of approximately 100s and 10s of SNPs due to current technological constraints, a preprocessing phase that heuristically split any arbitrarily large instance of SNPs into smaller chunk is necessary.

#### Supplementary Materials 8: Detailed analysis of the quantum computing experiments

In this section, we provide a detailed account of the experiments conducted on quantum devices. Firstly, we discuss the effectiveness of an optimization-based seeding procedure as compared to the random sampling-based procedure of Linear NeEDL. We then compare various quantum and classical devices in terms of execution speed to complete the optimization task. The primary objective of our experiments was to demonstrate that the optimization-based technique expedites the costly local search phase, which is based on a multitude of randomly selected seeds, by carefully selecting fewer seeds, namely those resulting from solving our optimization problem. Secondly, we investigate how the optimization process would scale on different devices with respect to the size of the dataset. Thirdly, we analyze the distribution of coefficients in the QUBO matrices to ensure their effective encoding on the quantum hardware without any loss of precision.

Thanks to the community detection tool, we are able to break any dataset into SNPs set of arbitrary size, allowing *quepistasis* to be tested on any kind of dataset. In principle, this allows us to solve datasets of arbitrary size on even the smallest quantum computer. However, we could use subsampling only for analyzing the quantum computing procedure scaling, while its use for quantum resource reduction, by working on smaller sets of data, was not viable due to the long waiting time on the quantum platform available on the cloud. In fact, for large sets, community detection generates a large number of subproblems, which, although very small in dimensions, must be sent to a quantum computer one by one, each contributing an overhead generated by the sending the task, waiting for the scheduling and receiving the results.

**Experimental setup**

To determine the potential speedup achieved through the quantum technique, we assessed the performance of NeEDL with and without the quantum computing module in operation.

Firstly, we performed NeEDL in a linear fashion without the use of quantum computing. In this scenario, the only requirement is to disable parallel execution and no specific configuration is necessary. Secondly, we evaluated the software with the quantum computer module enabled, which formulates the SNPs seeding problem as an optimization problem that can be then tackled using various algorithms. To achieve this, the community detection software should be configured to generate subsets of the SNPs seeding problem that are compatible with the selected optimization algorithm. The subsets should contain at least two SNPs and the maximum size can be arbitrarily selected. The optimization problem can be solved using several available algorithms, such as classical simulated annealing, which serves as the baseline classical technique, quantum annealing through D-Wave devices, and QAOA algorithm via a noiseless simulator and IBM superconducting-based quantum processing unit. The parameters utilized for solving each dataset are summarized in (Suppl. Table 14). Additionally, there are specific configuration parameters for each algorithm.

**Configuration of the thermal annealing**

The thermal annealing requires a random seed and specifies the number of output samples (i.e. how many times the annealing process is stopped and repeated), which is always set to 1000, as well as the number of sweeps (i.e. how many times the state of a solution is perturbed during the annealing process), which is always set to $10^5$. The annealing cooling schedule is not configurable and follows a geometric schedule, which is determined by the formula $T(i) = T(0) \cdot \alpha^i$. The initial values for this schedule are automatically selected based on the standard procedure implemented in the Ocean SDK software.

**Configuration of the quantum annealing**

The quantum annealing process involves several specific settings. Firstly, the number of output samples is set to 1000. The solver utilized is Advantage 6.1, which is the latest hardware available from D-Wave at the time of writing. The annealing operation is performed twice - the first time as a forward annealing and the second time as a reverse annealing. During the forward annealing, the process starts at time 0 with the Hamiltonian $H_I$, then, after $1\mu s$, it implements the Hamiltonian $0.5H_I + 0.5H_P$, pauses for $1\mu s$, and completes the annealing process after an additional $1\mu s$. During the reverse annealing, the process begins at time 0 with the state corresponding to the solution found at the end of the forward annealing, using the Hamiltonian $H_P$. It then goes back to $0.5H_I + 0.5H_P$ after $1\mu s$, pauses for $1\mu s$, and converges to $H_P$ again after another $1\mu s$. *quepistasis* allows to fully customize the quantum annealing schedule. It is worth noting that a longer schedule has the potential, in theory, to lead to better solutions by taking advantage of the adiabatic theorem. However, longer evolution periods also accumulate more noise on noisy devices, which can ultimately degrade the quality of the solutions. Therefore, it is generally more practical to increase the number of reads using shorter schedules rather than relying on a few reads with longer schedules. To utilize a

D-Wave device, one must either purchase the hardware or rent it and access it through the cloud. We opted for the cloud-based access provided through our partnership with CINECA.

##### Configuration of the QAOA algorithm

The QAOA algorithm involves several configurable settings. Specifically, the quantum circuit of QAOA is dependent on the number of repetitions of the scheme $e^{-i\beta H_B}e^{-i\gamma H_C}$. Its training process requires a specific selection of optimizers, which in our case is the stochastic gradient descent algorithm ADAM, and a maximum number of epochs for the training. Furthermore, the statistical error in the output of the quantum circuit relies on the number of shots. The QAOA can be simulated on a classical device for instances of quantum circuits having a small number of qubits. The simulation employed is a linear algebra-based procedure that is noiseless (except for the statistical error in the sampling of solutions). For our simulated QAOA experiments, we executed the QAOA ansatz ten times, with random initialization of each parameter drawn from a uniform distribution ranging from $[-2\pi, 2\pi]$. The circuit was trained for a maximum of ten epochs, although this number does not ensure convergence. Nonetheless, it is ten times larger than what can be accomplished in a realistic time frame using actual hardware, given that there are hundreds of independent subproblems to solve with QAOA for our datasets. We utilized 1024 shots. In order to use the IBM Perth and Lagos devices, one must subscribe to the IBM cloud-based service. Our partnership with CERN has provided us with access to this service. However, access to the machine is shared with a community of users, and requests are managed on a first-in, first-out (FIFO) basis, which can often lead to significant delays in obtaining results. In such case, we have repeated the QAOA ansatz once, and trained for 10, 5, and 1 number of epochs for datasets of 100, 500, and 1000 SNPs, respectively. The number of shots is 128. To utilize the IonQ Harmony device, it is required to have a subscription with the IonQ cloud-based provider. We have been granted access to this service through the Microsoft Azure Quantum platform. However, access to the IonQ device is also severely limited by a queue of users sharing the device. The QAOA algorithm software is based on the Qiskit library for Python, and it is made feasible to execute on both the simulator and quantum processing units through the use of various plug-ins.

##### Configuration of the QUBO formulation

There are some important considerations to be made about the parameters listed in (Suppl. Table 14). The value of parameter $K$ is consistently set to 5, indicating that the optimizer is directed to return SNP sets of approximately the same size. The strength of this constraint varies based on the relationship between the values of $\lambda_0$ and $\lambda_1, \lambda_2$. This implies that for subproblems of size 5 (used in QAOA experiments), the solution for the subproblem is expected to be the SNP set that corresponds to the entire subproblem. In such a case, the effectiveness of the approach depends almost entirely on the community detection tool. To minimize the size of each subproblem, the number of $N$ of SNP sets returned is always set to 1. There is a trade-off between the use of the community detection tool and the value of $N$. A larger value of $N$ increases the optimization problem size, necessitating a more aggressive community detection policy to compensate. Given more resources (qubits), this problem may be less significant. The interpretation of the $\nu$ parameters is related to the method of constructing weights for pairwise SNP correlation. Specifically, each weight between SNP $i$ and $j$ is determined using the formula $w_{i,j} = \nu w_{i,j}^{(1)} + (1-\nu)w_{i,j}^{(2)}$, where $w_{i,j}^{(1)}$ denotes the statistical correlation between the SNPs, and $w_{i,j}^{(2)}$ represents biological information.

##### Normalization of QAOA seeding time on IBM devices

To perform our experiments using IBM devices, we had to choose between renting the machine for a fixed amount of time or submitting circuits to a job queue for processing. However, the time spent in the queue is not an accurate reflection of the actual time required to complete the job, since it depends on the usage of the machine at that particular moment. To account for this, we calculated the average execution time of a quantum program (which includes one quantum circuit estimated with one or several sets of parameters). We then multiplied this average time by the number of subproblems in the dataset, along with the initial startup time, to obtain a more accurate estimate of the total time required. To calculate the average execution time, we excluded the top 25% highest and lowest scores from any job execution.

##### Comment on the performance analysis

An analysis of our experiments is shown in Suppl. Figure 8. As anticipated, the seeding procedure of Linear NeEDL is generally faster than the optimization-based approach since the generation of random seeds is extremely rapid. However, the two techniques are comparable in terms of their order of magnitude. The little advantage achieved by Linear NeEDL is quickly lost during the searching phase, which requires checking a vast number of random seeds and takes up to three orders of magnitude longer than the optimization procedure of *quepistasis* to complete. This is because the latter produces fewer seeds, which are obviously quicker to verify and produce solutions with either the same (BD, RA datasets) or even superior (T2D dataset) statistical scores.

Although different techniques often result in seeds having varying statistical scores (except for RA with 100 SNPs, see Suppl. Figure 8, near values $x = 0$), the local search technique consistently (heuristically) recovers the best outcomes (see Suppl. Figure 8, near values $x = 10^6$). The classical baseline thermal annealing algorithm demonstrates comparable performances with datasets that have a smaller number of SNPs. However, as the number of SNPs increases, this advantage quickly dissipates, and thermal annealing becomes the slowest technique. The simulated QAOA algorithm often produces the highest-quality initial seeds in terms of maximum likelihood, mainly due to the community detection tool. QAOA subproblem instances consist of 5 qubits (5 SNPs), and according to our formulation and configuration, we expect QAOA to return the entire set of 5 SNPs for each subproblem instance. However, the classical simulation of QAOA is computationally intensive, resulting in seeding times comparable to the classical baseline (for smaller sets, simulated QAOA is slower, while for larger ones, it is faster). The performance of QAOA executed on IBM devices closely mirrors that of simulated QAOA, even though the noise of these devices leads to suboptimal solutions, which negatively affects the local search execution time. The seeding time of the QAOA on the IBM device had to be normalized to avoid misleading results due to the long wait in the queue for access to the shared device on the cloud platform. The details of this process are shown in the next paragraph. Finally, the D-Wave device appears to struggle to find high-quality seeds in terms of statistical score, even though the local search can still recover the best results. This technique is often the fastest in terms of both seeding time and local search time: starting with the D-Wave seeds, the local search converges quickly to the best solution.

##### Comment on the scaling analysis

In Figure 5, we propose a scaling model for our approach to studying the seeding time of the optimization-based seeding procedure with respect to the dataset size. We conducted experiments on classical and quantum devices using subsamples of 100, 500, and 1000 SNPs for each of the three diseases tested. This enabled us to show the linear growth of seeding time, as evidenced by lines with higher slopes indicating better scaling. The classical baseline algorithm, thermal annealing, performed the worst. The results are followed closely by the QAOA approach on both simulated and IBM devices. The QUBO formulation constrains the algorithm to return cliques of at least size 5, and since this is also the maximum number of SNPs for a subproblem in the configuration we tested using QAOA, the algorithm often returns the entire set of SNPs for the subproblem, resulting in similar outcomes (although QAOA (IBM) results are noisy). The poor scaling of QAOA is due to the community detection tool having to split the problem into many small sub-instances to make it feasible on current IBM quantum devices. D-Wave devices exhibited the best scaling: due to the larger number of qubits, the community detection tool was able to create instances as large as thermal annealing and much larger than QAOA. With fewer subproblems, the technique took much less time. Lastly, it is important to notice that while linear scaling is appropriate for datasets of modest size, there is no guarantee that this trend will continue for significantly larger datasets.

##### Range and sparsity of QUBO coefficients

The distribution of coefficients is a crucial factor in solving a QUBO or Ising formulation, particularly with respect to sparsity. Sparse QUBO matrices require less overhead on the D-Wave device. Additionally, coefficients should be uniformly distributed since they are rescaled to a specific interval (e.g., $[-1, 1]$). A few coefficients with a significantly larger magnitude than most others can result in a concentration of values around zero, which could cause problems with encoding precision. To illustrate this point, we have presented the coefficient distribution in Suppl. Figure 14. As shown in the figure, it can be observed that the average standard deviation of $h$ and $J$ coefficients is, once rescaled, always above the encoding precision of the devices (less than $10^{-3}$ for gate-based quantum devices, less than $4 \times 10^{-3}$ for D-Wave Advantage device) and therefore we can assume the problem has been encoded with satisfactory precision on these devices.
